# Would Lifting Versus Maintaining COVID-19 Containment Policies Have Reduced Psychological Distress in the US?

**DOI:** 10.64898/2026.03.06.26347802

**Authors:** Mihael Cudic, Juan F de la Hoz, Lorenza Dall’Aglio, Justin D. Tubbs, Omid V. Ebrahimi, Emily Madsen, Daniel Fatori, Pedro F Zuccolo, James Lian, Dia Kabir, Yu Zhou, Devon Watts, Karmel W. Choi, Gisele G. Manfro, Ellen Sweeney, Yen-Feng Lin, Daisy Fancourt, COVID-19 Global Mental Health Consortium (CGMHC), Vikram Patel, Ronald C. Kessler, Sarah Bauermeister, Andre R Brunoni, Younga Heather Lee, Jordan W. Smoller

## Abstract

**Background:** Both the COVID-19 pandemic and containment policies caused widespread psychological distress, yet their independent effects remain unclear. Disentangling these effects could inform future responses that balance physical and mental health. This study sought to estimate the effect of lifting versus maintaining containment policies on psychological distress, independent of pandemic severity.

**Methods:** We conducted a state-level longitudinal analysis using the Behavioral Risk Factor Surveillance System (BRFSS), a representative survey of US adults, restricted to the pandemic period preceding widespread vaccine availability (April 2020 to April 2021). The exposure was lifting versus maintaining containment policies (school closures, workplace closures, event cancellations, and full lockdown) from the Oxford COVID-19 Government Response Tracker. Exposure was measured during periods of low (<25/100,000 new cases) or declining (>14 days) pandemic severity. The primary outcome was prevalence of psychological distress, derived from a BRFSS survey item corresponding to a PHQ-4 score ≥6.

**Findings:** Three causal inference approaches yielded consistent evidence of transient policy-lifting effects: (1) synthetic control analysis of Maine showed a temporary 5.5 percentage-point reduction in psychological distress lasting three months before returning to counterfactual levels; (2) within-state fixed effects found immediately after lifting full lockdown, distress decreased by 5.68 [-8.67, −2.69] percentage points, declining by 30 days (−3.24 [-6.88, 0.39]) and negligible at 60 days (−0.94 [-3.77, 1.89]); (3) target trial emulation detected no significant effects from lifting versus maintaining policies for 90 days.

**Interpretation:** Lifting containment policies in the first year of the pandemic produced immediate but transient reductions in psychological distress. These results suggest that extended containment policies were unlikely to account for persistent increases in distress during this period.

## INTRODUCTION

COVID-19 containment policies led to widespread disruptions of daily life, with a growing body of evidence documenting their detrimental effects on mental health ^1–15^. Evidence from longitudinal studies ^16,17^ and systematic reviews ^18^ suggests that stricter lockdowns were associated with increases in psychological distress, especially among specific subpopulations such as young adults ^19^, those with pre-existing mental health conditions ^1^, and medical professionals ^20^. However, it remains unclear to what extent these mental health outcomes are attributable to containment policies beyond other pandemic-related stressors (e.g., infections and deaths in the community, economic uncertainty) ^6–9^.

Isolating the effects of the containment policies remains methodologically challenging primarily due to *confounding by indication*: containment policies were implemented in response to pandemic severity, which itself could affect mental health. Additionally, the near-universal implementation of containment policies during the early pandemic (i.e., school, workplace, and public event closures) limits the ability to estimate average treatment effects due to lack of counterfactual comparisons. Here, by examining variation in the timing of policy lifting during periods of low or declining pandemic severity, we address both challenges: reducing confounding and providing variation for causal contrasts.

Other aspects of several prior studies have also limited causal inference and interpretability, including reliance on aggregate measures of policy stringency that obscure the effects of specific policies ^13,14,21^ and the use of convenience samples and cross-sectional designs that preclude assessment of prospective relationships between policy changes and mental health ^11,15,22^. In contrast, we examine the effects of lifting versus maintaining *specific* containment policies on mental health using three longitudinal, nationally representative data sources and complementary causal inference designs.

To facilitate causal inference in the highly complex setting of the pandemic, we apply a triangulation strategy using three complementary quasi-experimental designs. First, using synthetic control (SC) ^23^ analyses, we estimate the counterfactual trajectories for a state that lifted containment policies early, using weighted combinations of comparable states that maintained policies. Next, our within-state fixed effects (WSFE) models compare outcomes before and after policy changes within the same state, controlling for time-invariant confounders. Finally, our target trial emulation (TTE) estimates the average treatment effect of lifting versus maintaining containment policies for 90 days, emulating a randomized controlled trial with observational data ^24^.

## METHODS

### Data Source and Study Population

We compiled longitudinal data on all 50 US states from multiple sources. Systematic records of state-level pandemic interventions from 2020-2022 were obtained from The Oxford COVID-19 Government Response Tracker (OxCGRT ^25^), which documents government containment policies, economic support, and vaccination uptake. Daily COVID-19 case and death counts for each state, spanning January 2020 to March 2023, were gathered from The New York Times COVID-19 Data Repository ^26^. Mental health data was extracted from The Behavioral Risk Factor Surveillance System (BRFSS ^27^), an annual telephone survey of over 400,000 US adults tracking health behaviors and conditions. Finally, state-level sociodemographic data were sourced from The US Census Bureau ^28^.

### Pandemic Timeline

The US White House declared a national emergency on March 13, 2020 ^29^. The next year, on May 13, 2021, the US Centers for Disease Control and Prevention (CDC) issued guidance stating that fully vaccinated individuals no longer needed to “physical distance in any setting” except where legally required ^30^. Our analysis focuses on this pre-vaccination guidance period of the pandemic, which we defined as April 1, 2020, to April 30, 2021 (Figure 1). This timeframe captures the complete months when containment policies remained the primary pandemic response tool and excludes the initial weeks of crisis response and rapid policy implementation.

**Figure 1.**
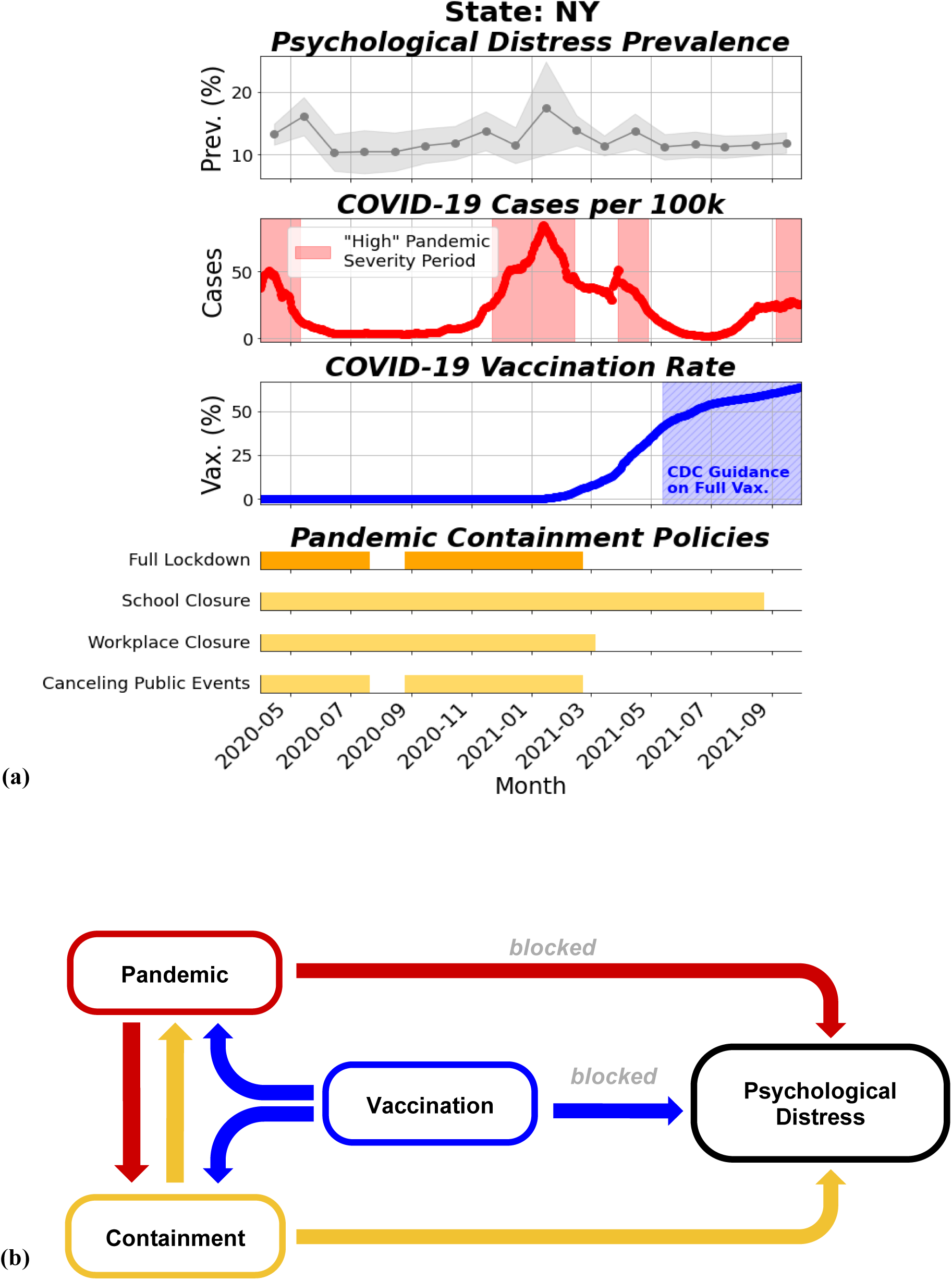
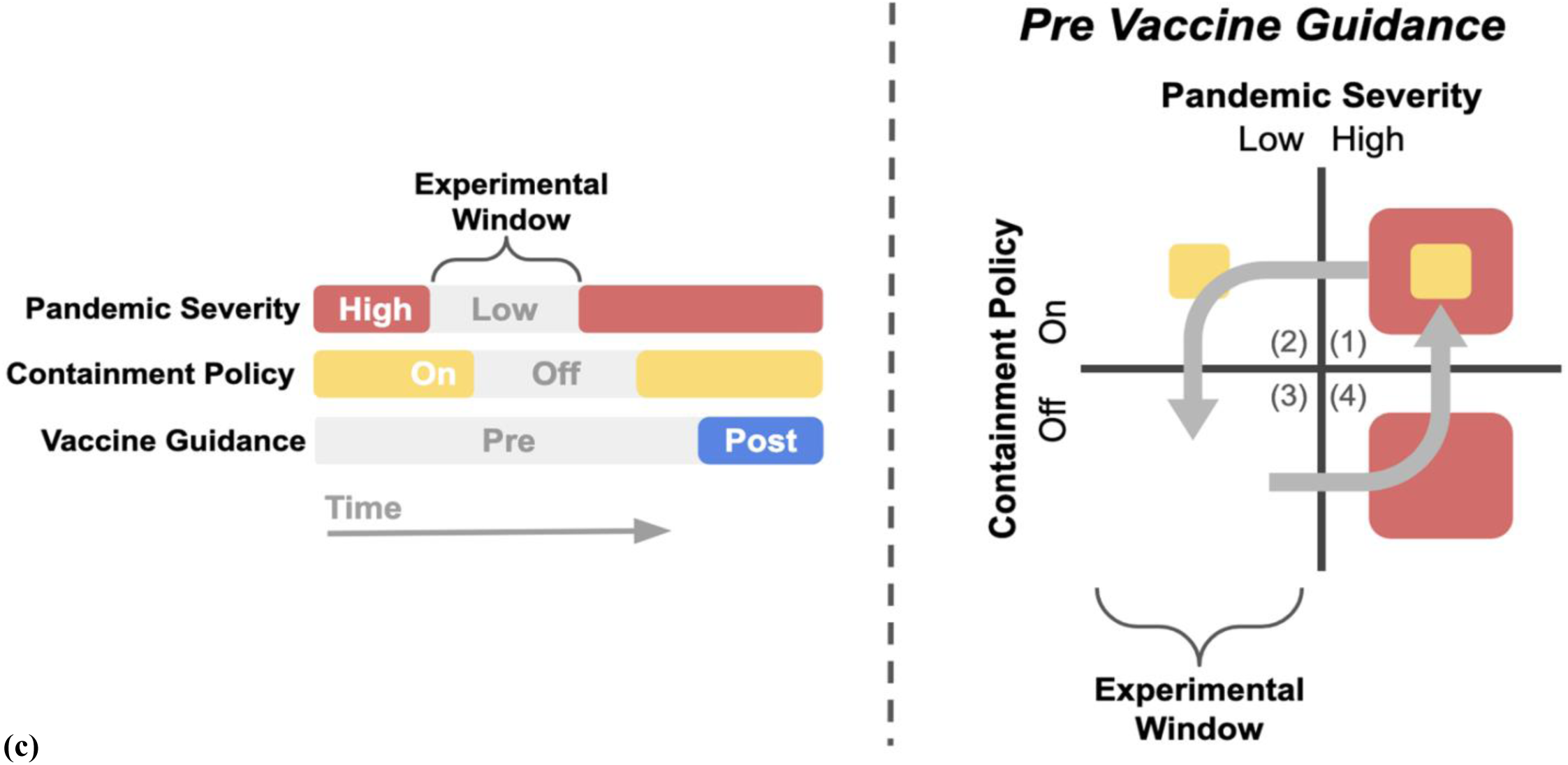
Pandemic severity, policy implementation, and psychological distress: observed patterns and conceptual frameworks to mitigate confounding by indication. **(a)** A visualization of four pandemic-related indicators in New York from April 2020 to April 2021 is shown. Top: Monthly prevalence of psychological distress, with 95% confidence intervals (shading). Estimates represent the percentage of individuals reporting more than 14 days of poor mental health in the past 30 days, based on BRFSS survey responses. Upper Middle: Daily COVID-19 case rates from NYTCDR. Time periods are classified as “High” or “Low or declining” pandemic severity based on rate trends. Lower Middle: Vaccine uptake data from OxCGRT. Notably, the CDC issued guidance for fully vaccinated individuals on May 13, 2021. Bottom: Timeline of three mandated containment policies—school closures, workplace closures, and public event cancellations—sourced from OxCGRT. Days when all three were simultaneously active are marked as full lockdowns. **(b)** A conceptual illustration of confounding by indication during the pandemic, where containment policies are typically enacted during periods of “high” pandemic severity. Vaccine uptake also confounds associations, making it difficult to isolate the effects of policies on psychological distress. Our study is designed to block direct paths from pandemic severity and vaccination to psychological distress. **(c)** Our conceptual framework to mitigate confounding. Specifically, we envision four experimental conditions: (1) “high” pandemic severity with active policies, (2) “low or declining” pandemic severity with active policies, (3) “low or declining” pandemic severity without policies, and (4) “high” pandemic severity without policies. Our analyses focus on conditions (2) and (3), that is, periods of low or declining pandemic severity occurring before vaccine guidance, when the independence of containment policy activity case rates was maximal.

### Measurements

#### Exposures

We focused on three containment policies tracked by OxCGRT: school closures, workplace closures, and cancellation of public events. We selected these policies because: a) they were adopted by nearly all US states at various times throughout the pandemic, and b) implementation varied in whether they were required versus recommended (eMethods 1, eFigure 1). OxCGRT codes the stringency of these policies as 0-no policy, 1-recommendation, 2 or 3-partial or full requirement. We then binarized them as absent (0–1) or present (2+). In addition to individual policies, we examined “full lockdown”, which we defined as the simultaneous implementation of all three policies. Periods with only one or two active policies were classified as “partial lockdown.”

#### Pandemic Severity

We classified each day of the pre-vaccination guidance period (April 2020 to April 2021) as belonging to a “low or declining” or a “high” severity period using state-level COVID-19 case trends from the NYTCDR. “Low or declining” periods began when cases either fell below 25 per 100,000 (CDC ^30,31^ and European thresholds ^32^) or declined for 14 consecutive days (The White House reopening guidelines ^33^). These periods continued until cases spiked in a single day or increased for a week, exceeding 25 per 100,000. All remaining time was classified as “high” severity periods (Figure 1a). Sensitivity analyses of severity classification parameters showed that thresholds for minimum case rate had the greatest impact on the classification (eFigure 2). To assess robustness of our findings, we therefore present additional analyses using 10 and 50 cases per 100,000 thresholds, the lower and upper bounds of moderate case load per CDC definitions.^33^

**Figure 2.**
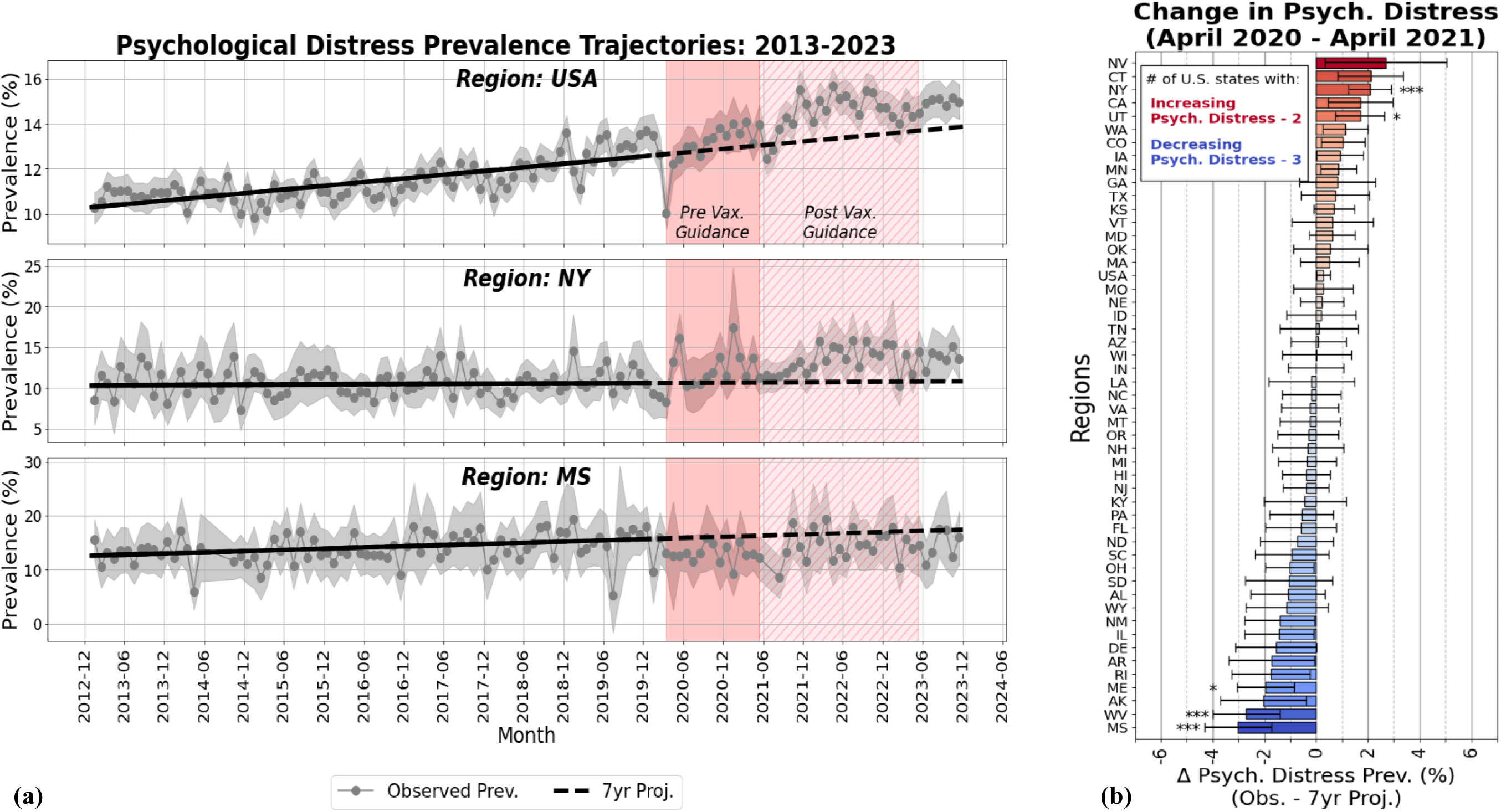
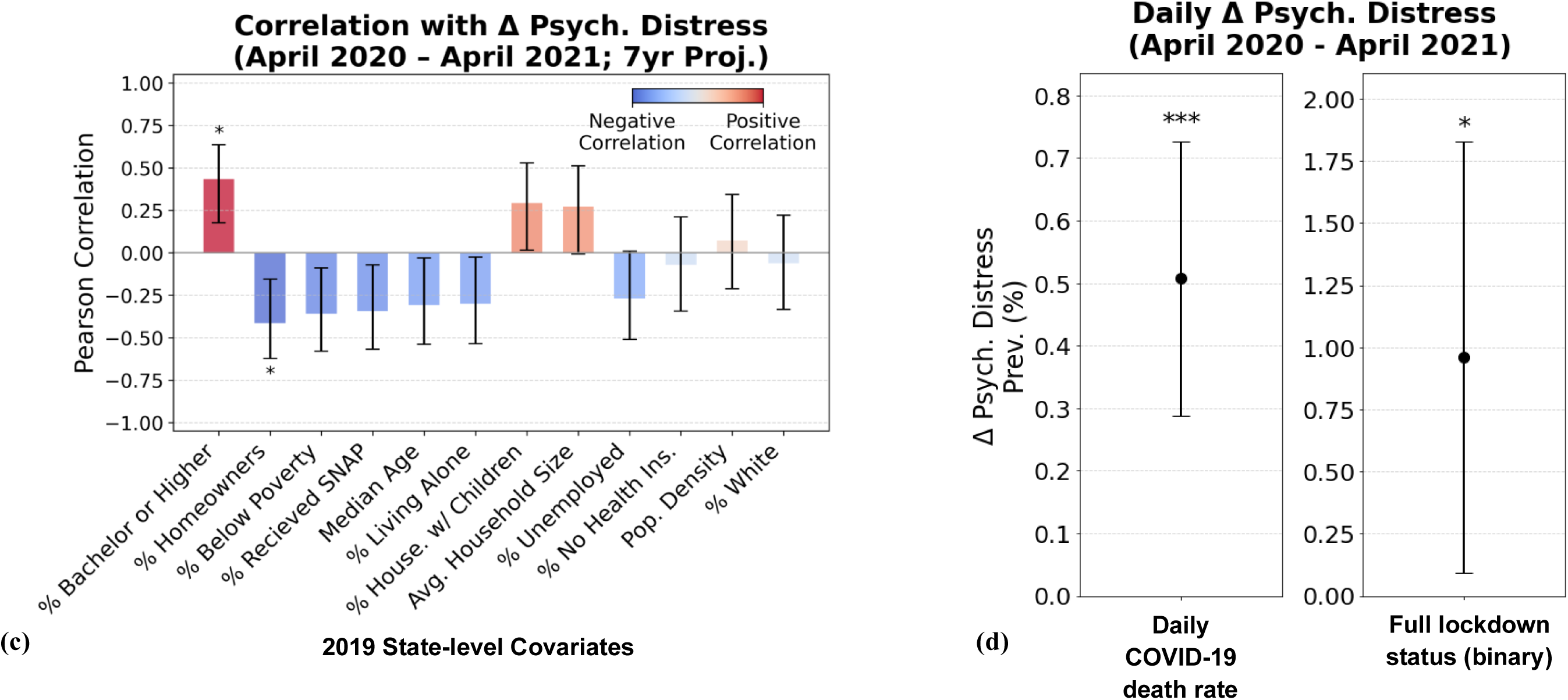
Regional trends and correlates of psychological distress before and before and during the COVID-19 pandemic. **(a)** Estimated psychological distress prevalence trends for the US, New York, and Mississippi from 2013 to 2023. Fitted 7-year pre-pandemic trends (2013–2019) and their projections through 2023 are shown as solid and dashed black lines, respectively. The pandemic period is divided into two stages, pre- (dark pink shading) and post-CDC guidance (light pink shading) for fully vaccinated individuals (March 13, 2020 to May 12, 2021, and May 13, 2021 to May 11, 2023). **(b)** Total change in psychological distress (observed minus 7-year projected prevalence) from April 2020 to April 2021 across US states and nationwide. Significance thresholds: p < 0.05/51 *, p < 0.01/51 **, p < 0.001/51 ***. **(c)** Pearson correlation of distress outcomes (from panel b) with 2019 state-level covariates sourced from the US Census. Significance threshold: * p < 0.05/12 **(d)** The associated daily change in psychological distress prevalence from daily COVID-19 deaths per 100,000 and an indicator for full lockdown status (1 = fully locked down, 0 = no policy in place, i.e., partial lockdown days were omitted in the analysis) from April 2020 through April 2021, controlling for month and state fixed effects. Significance thresholds: p < 0.05 *, p < 0.001 ***. 95% confidence intervals are displayed for every subfigure.

#### Outcomes

Psychological distress was measured using question Q2.2 of the BRFSS (“Now thinking about your mental health, which includes stress, depression, and problems with emotions, for how many days during the past 30 days was your mental health not good?”). Survey responses were dichotomized into not psychologically distressed (0 −14 days) and psychologically distressed (>14 days), which has been shown to correspond to a score of 6 or greater on the 4-item Patient Health Questionnaire (PHQ-4; AUC=0.84; false positive rate = 7.2%; false negative rate = 6.2%) ^34^ and aligns with the duration criteria for depression in the DSM-5 ^35^. For any given US state and time period, we estimated psychological distress prevalence by calculating the weighted proportion of respondents classified as distressed, using BRFSS survey weights and accounting for different survey years (eMethods 2). Standard errors for prevalence estimates were calculated using Taylor series linearization, as implemented in the survey R package ^36^. For our results, we report the absolute change in prevalence (i.e., change in percentage points).

#### Covariates

State-level social, economic, housing, and demographic characteristics were sourced from the 2019 Census Bureau data. From the complete set of available state-level indicators, 12 variables were chosen: percentages of adults with a bachelor’s degree or higher, individuals living alone, households with children, residents without health insurance, and those identifying as white; rates of homeownership, poverty, unemployment, and SNAP participation; averages for median age, household size; and population density. These indicators were chosen based on prior research identifying their relevance as determinants of mental health outcomes during the COVID-19 pandemic.^34^

### Statistical Analysis

#### Overview of Analytic Strategy

We started by characterizing secular trends in psychological distress throughout the pre-vaccination guidance period (April 2020 to April 2021), then estimated causal effects of lifting containment policies (full lockdown, school closures, workplace closures, cancellations of public events) using three complementary approaches designed to isolate policy effects from pandemic severity. First, we used Maine, the first state to lift its full lockdown, as a case study and applied SC analysis to estimate the counterfactual trajectory in which Maine delayed lifting its lockdown to match the timing observed in a weighted combination of comparable states. Second, we applied within-state fixed effects (WSFE) analysis to assess the average effect of lifting policies during “low or declining” severity periods across US states. Third, we implemented TTE to estimate the average treatment effect of immediately lifting versus maintaining containment policies for 90 days when transitioning from “high” to “low or declining” severity periods (eMethods 1-6 for detailed description of each analytic approach). All analytic code can be found in the associated GitHub repository (https://github.com/cgmhc/Target-Trial-Emulation).

#### Secular Trends of Psychological Distress

We estimated changes in psychological distress prevalence by comparing observed prevalence during the pre-vaccination guidance period with expected prevalence projected from 7-year pre-pandemic trends (January 2013 to December 2019) using weighted linear regression (Figure 2, eMethods 3). We additionally examined correlations between prevalence changes and 2019 state-level sociodemographic characteristics, and tested associations of daily distress with COVID-19 deaths and full lockdown implementation using state and month fixed effects models (eMethods 3).

#### Synthetic Control (SC) Analysis – Maine

We used SC methods to estimate the counterfactual trend in psychological distress for Maine. Maine was selected as the target state, as it was the first state to lift its full lockdown during a period of low or declining pandemic severity on September 14, 2020 (Figure 3a, eMethods 4). Using 7 years of pre-intervention data, we identified a set of four comparable states that maintained full lockdowns for at least five additional months (Washington, New Mexico, Oregon, and California) and constructed a synthetic control as a weighted combination of these states (eFigure 8).

**Figure 3:**
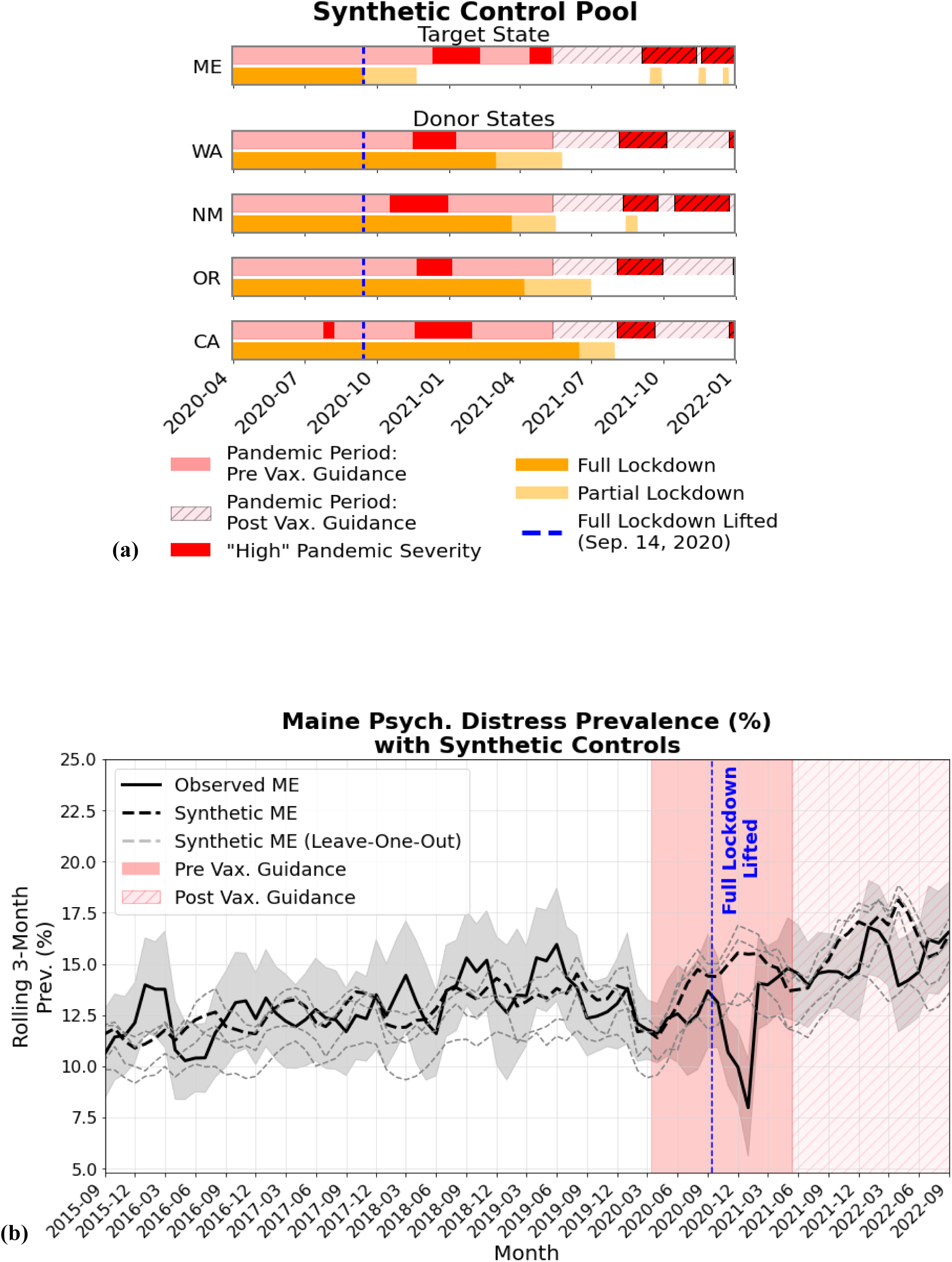
Estimating the impact of lifting full lockdown early in Maine using synthetic controls. **(a)** Timeline showing periods of “high” pandemic severity and full lockdown status in Maine, along with four donor states (Washington, New Mexico, California, and Oregon) used to generate synthetic controls. “High” pandemic severity periods appear in solid red before CDC guidance for fully vaccinated individuals (May 13, 2021) and hatched red afterward. Full lockdown periods are indicated in orange and partial lockdown periods in light orange. The dashed blue line marks the date Maine lifted its full lockdown (September 14, 2020). **(b)** Trends in Maine’s observed psychological distress prevalence (solid black line with 95% confidence intervals) and estimated prevalence trends from the counterfactual scenario had Maine lifted all policies after vaccine guidance (dashed black). Prevalence estimates were reported monthly, with each value representing a 3-month centered average to reduce noise in monthly fluctuations. The counterfactual was estimated using synthetic control analysis, creating a weighted average of distress prevalence from the four donor states shown in panel **(a)**. Weights were calibrated using data from January 2013 to August 2020 (WA 0.39, NM 0.32, CA 0.19, OR 0.15) and fixed thereafter to project Maine’s counterfactual trend. Gray dashed lines indicate leave-one-out counterfactual estimates, serving as a sensitivity analysis (eMethods 4).

### Within-State Fixed Effects (WSFE) Analysis

We examined time-varying effects of lifting containment policies using WSFE models that leverage within-state changes in policy status over time. Specifically, we compared psychological distress prevalence in 30-day windows before and after policy changes, with washout periods of 0, 30, and 60 days to assess the persistence of effects (Figure 4a, eMethods 5). Analyses were restricted to periods of low or declining pandemic severity for all four policies. Models additionally included time-varying covariates for prior policy duration and other concurrent policies.

**Figure 4.**
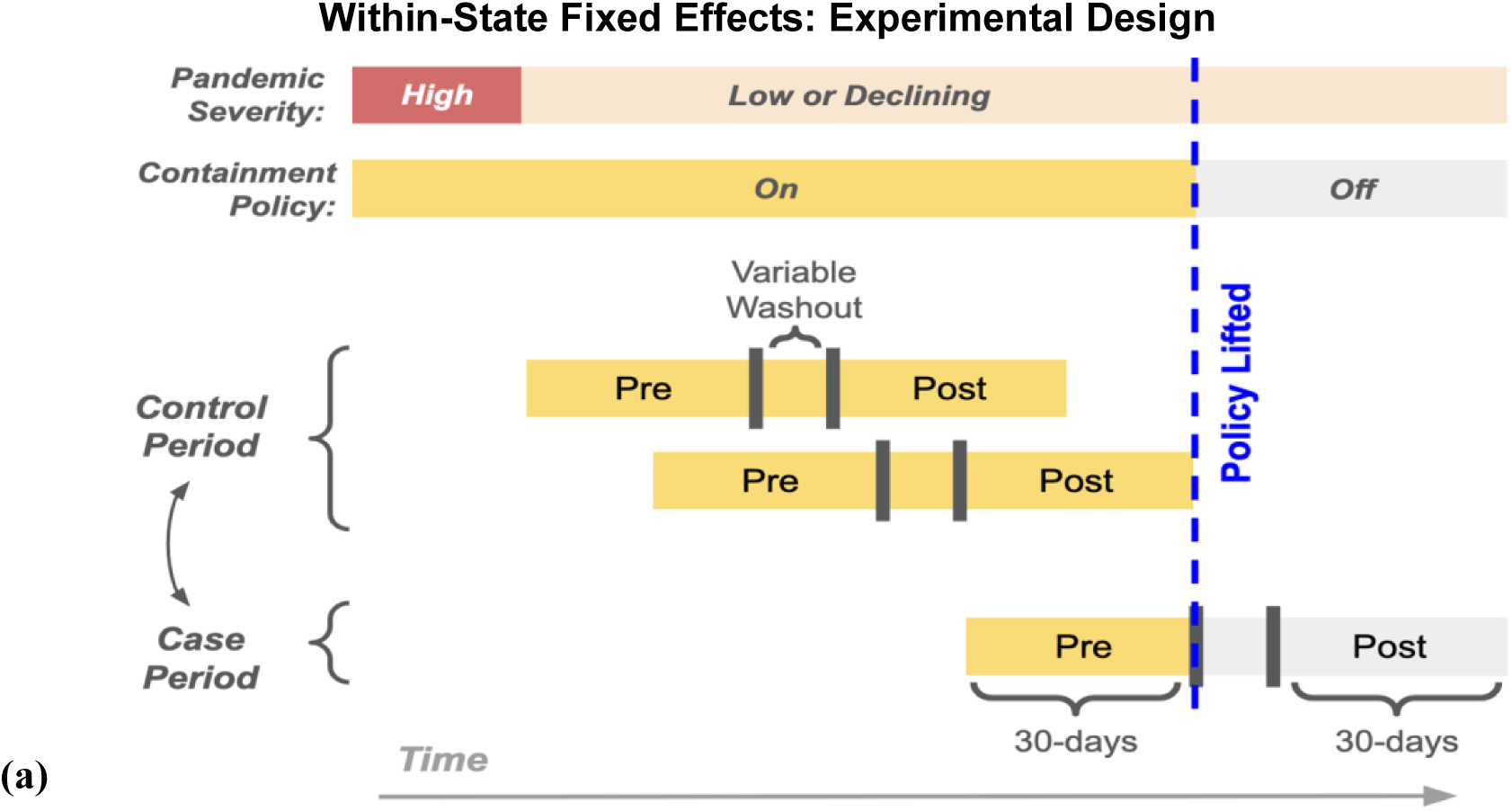
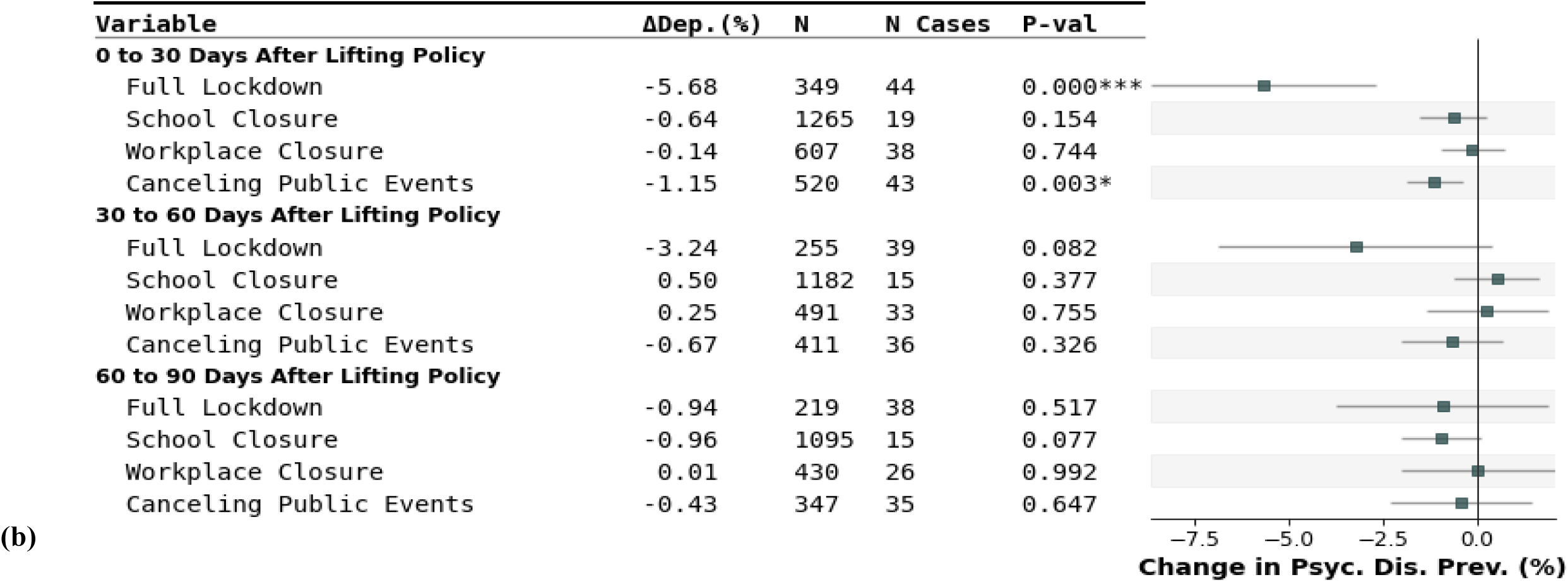
Estimating the short and long-term effect of lifting policies on psychological distress using within-state fixed effect models. **(a)** Experimental design for the within-state fixed effects analysis. States contributed “control periods”, when policies were continuously maintained, and “case periods”, when policies were lifted (i.e., treated) after 30 days and remained lifted. Eligible observations were limited to periods where the full 30-day pre-treatment window occurred when the pandemic was “low or declining” and before the CDC’s May 13, 2021 guidance for fully vaccinated individuals. The effect of policy lifting was estimated by comparing pre- and post-treatment 30-day prevalence within both case and control periods. Variable washout periods applied in separate experiments to assess the persistence of the treatment effect. **(b)** Estimated effects of lifting four types of policies—including full lockdown, school closures, workplace closures, and cancellation of public events—on 30-day psychological distress prevalence, compared to a counterfactual scenario in which policies remained in place. Effects were evaluated at 0, 30, and 60 days following each policy change. Estimates were obtained using weighted linear regression models, regressing post-treatment prevalence on treatment status, while adjusting for pre-treatment prevalence, month and state fixed effects, and time-varying confounders (eMethod 5). Significance thresholds: p < 0.05/4 *, p < 0.01/4 **.

#### Target Trial Emulation (TTE) Analysis

We emulated a target trial to estimate the effect of lifting versus maintaining containment policies on psychological distress over a 90-day exposure window, with all four policies evaluated independently (Figure 5a, eMethods 6). This approach compared sustained policy strategies defined at baseline among state-trials that became eligible when pandemic severity transitioned to low or declining following a 30-day run-in period with an active policy. Average treatment effects were estimated using parametric G-computation and inverse probability weighting for censoring ^37–39^. Models were adjusted for 2019 state-level covariates (education and homeownership), days since the pandemic began (March 13, 2020), and a binary indicator for whether the policy remained beyond 14 days in the follow-up window (eTable 1).

**Figure 5.**
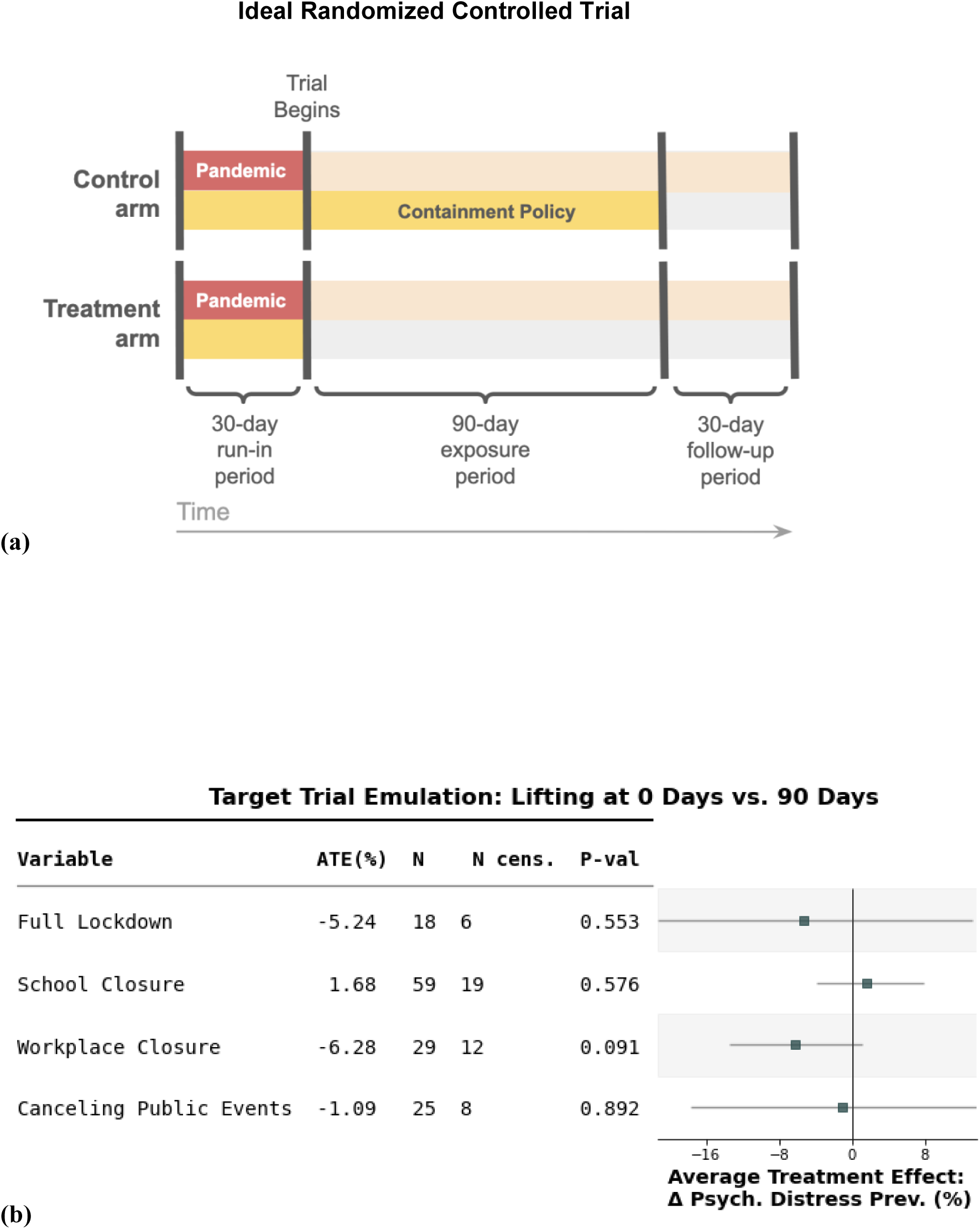
Emulating a target trial to estimate the impact of early policy lifting on psychological distress. **(a)** A diagram illustrating the design of an ideal randomized controlled trial (RCT) used to emulate our target trial. The design includes: (1) a 30-day run-in period with both “high” pandemic severity and active policy measures to determine eligibility; (2) a 90-day exposure window; and (3) a 30-day follow-up period to assess the change in prevalence. Within each emulated state-level trial, states were assigned to either a control arm, where policies were maintained throughout the 90-day exposure window, or a treatment arm, where policies were lifted immediately at the trial’s start. The trial’s outcome was defined as the change in psychological distress prevalence from the run-in period to the follow-up period. Details on how the target trial was adapted for our emulation are provided in eMethod 6, eTable 1. **(b)** Estimated average treatment effects (ATE) and bootstrapped 95% confidence intervals of a 90-day intervention involving full lockdown, school closures, workplace closures, and cancellation of public events. Estimates were obtained using parametric g-computation with inverse probability weighting to account for censorship. Models were further adjusted for key state-level sociodemographic covariates.

## RESULTS

### National and State Changes in Psychological Distress

From April 2020 through April 2021, the absolute increase in psychological distress prevalence among US adults was 0.28 percentage points (95% CI: 0.02-0.54; p=3.4e-2) above the expected 7-year pre-pandemic trend (Figure 2a). In our sensitivity analysis, estimates of prevalence change varied by baseline period: 0.25 percentage point decrease (95% CI: −0.53 to 0.00) using a 5-year trend, and 0.86 percentage point increase (95% CI: 0.60-1.12) using a 9-year trend. When national estimates were disaggregated, substantial variation emerged across US states (Figure 2b, eFigure 6a-6b). Only New York (increase) and Mississippi (decrease) consistently had statistically significant changes across all baseline trends (5-, 7-, and 9-year trends). Notably, Hawaii showed no significant change despite having one of the strictest containment policies, experiencing the lowest COVID-19 case and death rates, and the fewest high-severity days between August 2020 and April 2021 (eFigure 7).

### Sociodemographic and Pandemic Predictors of Psychological Distress

We identified two significant state-level sociodemographic predictors of psychological distress change: education and homeownership (Figure 2c, eFigures 6c-d). States with higher proportions of residents holding bachelor’s degrees showed increased distress (r=0.43; 95% CI: 0.18-0.64; p=1.7e-3), while states with higher homeownership rates showed decreased distress (r=-0.42; 95% CI: −0.62 to −0.16; p=2.7e-3). These associations remained robust using different baseline periods (5-, 7-, and 9-year trends).

Daily COVID-19 death rates and concurrent implementation of lockdown policies, including school closures, workplace closures, and cancellation of public events, were also significantly associated with psychological distress (Figure 2d). Each additional death per 100,000 was associated with a 0.51 percentage points increase in distress prevalence (95% CI: 0.29-0.73; p=5.5e-6), while each day under full lockdown was associated with a 0.96 percentage points increase (95% CI: 0.09-1.83; p=0.030).

### SC Analysis – Maine

Maine’s psychological distress prevalence was 12.5% (95% CI: 9.8-15.2) in July to September 2020 (Figure 3). Following the end of full lockdown on September 14, 2020, distress declined to 10.0% (95% CI: 8.2-11.7) in November 2020 to January 2021, a decrease of 2.5 percentage points. It then rose to 14.0% (95% CI: 12.1-15.9) in February to April 2021, slightly exceeding pre-lockdown levels.

The SC had a good pre-intervention fit (R² = 0.28). In November 2020 to January 2021, the counterfactual prevalence estimate was 15.5%, significantly higher than Maine’s observed 10.0% (difference: 5.5 percentage points; 95% CI: 2.6-8.5; p=1.5e-4). Following vaccine guidance in May 2021, Maine’s observed prevalence converged with the synthetic controls for the following two years, with no statistically significant monthly differences. Sensitivity analyses excluding Maine (i.e., leave-one-out) and varying training window durations confirmed robustness to donor composition (eFigure 8). In summary, Maine experienced a short-term (3-month) reduction in psychological distress after lifting full lockdown, with distress levels returning to the counterfactual trajectory within five months.

### WSFE Analysis

Lifting full lockdown (44 case and 305 control periods across 45 states) produced an immediate and significant reduction in psychological distress (decrease of 5.68 percentage points, 95% CI: −8.67 to - 2.69; p=2.4e-4) (Figure 4b). As the washout length (i.e., days between pre and post treatment windows) increased, effects diminished, potentially due to fewer eligible case and control periods, and were no longer significant by 30 days (−3.24 percentage points, 95% CI: −6.88 to 0.39; p=8.2e-2). Canceling public events (43 case/477 control periods across 47 states) showed a similar pattern with a decrease of 1.15 percentage points with no washout period (95% CI: −1.89 to −0.41; p=2.6e-3) that did not persist with longer washouts. School closures (19 case/1,246 control periods across 50 states) and workplace closures (38 case/569 control periods across 50 states) showed no significant effects on distress for any washout period. Results were robust to alternative “high” and “low or declining” pandemic severity periods (eFigure 9). These findings suggest that across states, lifting some containment policies produced immediate but transient reductions in psychological distress, with effects dissipating within 30-60 days.

### TTE Analysis

We conducted four TTEs examining the effects of immediate lifting versus maintaining containment policies for an additional 90 days when pandemic severity transitioned from “high” to “low or declining” (eTable 1). For each treatment, we identified varying numbers of eligible US state-trials: full lockdown (18 state-trials, 6 censored; eFigure 10), school closures (59 state-trials, 19 censored), workplace closures (29 state-trials, 12 censored), and cancellation of public events (25 state-trials, 8 censored) (Figure 5). Across all four TTEs, the observed distribution of the policy duration aligned with our predefined exposure definition, with over half of all state-trials having either short (≤20 days) or long (≥70 days) exposure durations (eFigure 11). This bimodal distribution reflects clear separation between lifted and sustained policy strategies, supporting the interpretability of the 90-day exposure contrast (full lockdown: 50.0%, school closure: 62.7%, workplace closure: 62.1%, public event cancellation: 52.0%). Our power analysis showed that we would have 80% power to detect a change of approximately 4.25 percentage points with a sample size as small as 20 state-trials and a change of approximately 2.35 percentage points with a sample size of 60, respectively (eFigure 12).

Our G-computation models to counterfactually predict psychological distress showed reasonable model fit for most containment policies (R² = 0.28 for full lockdown, 0.27 for workplace closures, 0.22 for cancellation of public events), except for school closures (R² = 0.04). The low model fit for school closure likely stems from higher censoring and limited variability, since 34% of state-trials extended beyond the 90-day intervention window (eFigure 11), suggesting that school closure policies operated under different decision-making frameworks than other containment measures. Our findings show that, during the transition from “high” to “low or declining” pandemic severity, 90 days of no containment policies (i.e., “lifting”) versus 90 days of sustained containment policies showed no significant differences in psychological distress (Figure 5), including across all sensitivity analyses (eFigure 13).

## DISCUSSION

Understanding the mental health impact of lifting containment policies could help inform future pandemic responses. We applied three complementary analytic approaches to isolate these effects from those of pandemic severity. Among US adults, our findings suggest that lifting COVID-19 containment policies during periods of low or declining pandemic severity produced substantial but transient reductions in psychological distress that dissipated within 30 to 60 days. We observed this pattern consistently across three complementary methods designed to isolate policy effects from those of pandemic severity. Specifically, an analysis of Maine versus synthetic control states showed an absolute decrease of 5.5 percentage points in psychological distress prevalence lasting approximately three months. The WSFE models across all US states confirmed immediate reductions of 5.7 percentage points immediately after lifting full lockdown, with effects diminishing by 30 days and largely absent by 60 days. Finally, our TTE found no evidence for significant effects of maintaining policies for 90 days versus immediate lifting. Despite these estimated effects of policy lifting, national psychological distress prevalence increased only modestly during the first 13 months of the pandemic (April 2020 to April 2021), with state variations perhaps better explained by demographic and economic factors than the policies considered here.

Prior research on the psychological effects of containment policies has yielded mixed findings. Some studies documented increased distress during lockdown periods ^17,40^, while others reported modest or null differences after adjusting for pandemic severity ^4,18^. These inconsistencies reflect well-known methodological challenges, particularly the difficulty of isolating policy effects from broader pandemic dynamics ^41^. Studies addressing this challenge using modern methods designed to make causal inferences from observational studies, such as differences-in-differences, remain rare. Ferwana and Varshney ^1^, using US medical claims data, found that mental health tracked more closely with policy shifts than with infection trends. Similarly, Serrano-Alarcón et al. ^42^ found that, in Scotland and England, lifting stay-at-home orders led to a nominally significant improvement in mental health symptoms that dissipated within a month. Our study extends these findings by examining policy lifting effects across US states, providing actionable insights for policy decisions.

Our finding that reductions in psychological distress following policy lifting were short-lived may help contextualize discrepancies in the literature, in which studies with short follow-up windows often report larger effects, while longer-term studies tend to show attenuation or recovery ^41,43,44^. Although our study was not designed to test psychological adaptation explicitly, our results are consistent with longitudinal studies demonstrating time-varying effects of containment policies on psychological distress. Prior studies of policy implementation or maintenance observed attenuation of adverse effects on psychological distress over time^12,16,45,46^. In contrast, our analysis examined policy removal and identified a parallel transient response, with psychological distress improving immediately after lifting containment policies and returning to pre-intervention levels within one to two months.

Importantly, concurrent implementation of school closures, workplace closures, and cancellation of public events had measurable effects on psychological distress, whereas individual components had limited impact. If true, this may reflect a threshold or cumulative effect of policies, in which changes in psychological distress emerge only when multiple domains of daily life are simultaneously restricted or eased. Alternatively, this pattern might be attributed to the challenge of isolating the effects of individual policies, as they were often implemented together throughout the pandemic.

Finally, although our results indicate that lifting containment policies was associated with immediate changes in psychological distress within individual states, national prevalence increased by only 0.28% during the pre-vaccination guidance period, compared with a larger increase of 1.29% during the subsequent two years, when most containment measures had already been lifted (eFigure 1). Consistent with this pattern, Kessler et al.^34^ suggested that modest population-level changes observed early in the pandemic likely reflect underlying heterogeneity, with mental health impacts concentrated among specific subpopulations rather than uniformly distributed across the population. As a result, differences in reported mental health changes across studies may reflect variation in population composition as well as the timeframe under analysis.

Our results also suggest that states with higher levels of educational attainment experienced greater increase in distress, whereas those with higher homeownership rates saw less or even decreasing psychological distress. The link between higher education and increased distress appears counterintuitive, but reflects the mixed findings in the literature, with some studies showing worse outcomes among those with lower education ^47,48^ while others report greater increases in anxiety and depression among those with higher education and income ^49,50^. Homeownership, however, may act as a proxy for financial security and physical assets, both of which have been shown to buffer against psychological distress ^51^. Our findings align with previous work indicating a potential role of socioeconomic context in shaping mental health trajectories during the pandemic ^52,53^.

### Strengths

Our study triangulates evidence to examine an underexplored question for pandemic preparedness. Unlike prior research which has largely focused on policy implementation or policy stringency, we examine whether a timely lifting of containment policies in response to decreasing pandemic severity would have improved mental health. This focus on policy removal, rather than implementation, is particularly important since every US state eventually lifted policies, making our intervention broadly relevant and enabling the construction of plausible counterfactuals.

We further strengthen causal inference through several study design choices. First, we mitigate confounding by indication by restricting analyses to low or declining pandemic periods, when policy decisions are less directly tied to immediate threats to public health. Second, we limit our timeframe to the pre-vaccine guidance period, when non-pharmacological interventions were the primary tools for viral containment. Third, we utilize the BRFSS, a nationally representative longitudinal dataset that enables prospective assessment of policy effects across US populations. In addition to these key study design decisions, we addressed additional sources of confounding, including regional characteristics, time-varying factors, and pre-existing trends in mental health. By isolating policy effects in a single treated unit through SC analyses, controlling for time-invariant confounding through WSFE analyses, and emulating randomized assignment through TTE, we provide convergent evidence on the psychological impact of lifting containment policies during periods when such interventions were epidemiologically indicated.

### Limitations

Several limitations warrant consideration in interpreting our findings. First, our state-level ecological design constrained sample size, limiting statistical power to examine a broader set of confounders or explore treatment effect heterogeneity. Future studies could apply this framework at county or city levels, or expand internationally, to increase sample size, variation in policy implementation, and relevance to international contexts. Second, the validity of our analyses depends on several assumptions that may be challenged, including no unmeasured confounding (difficult to ensure with limited state-level and time-varying covariates), no interference between states (policies and outcomes may have spillover effects), and consistent treatment definitions across heterogeneous contexts (identical restrictions may have varied in enforcement, compliance, and practical impact across states).

Third, our exposure and outcome measurement presents additional challenges. The dichotomized psychological distress measure relies on self-reported surveys that may not reflect clinically meaningful outcomes or healthcare usage. As a result, we can only assess population-level prevalence changes within fixed time windows, potentially missing more granular temporal dynamics. Similarly, our binary classification of pandemic severity (high versus low or declining) was necessary for analysis but may not align with multilevel frameworks (e.g., low/medium/high risk) that were also used for policy decisions ^54–56^, potentially creating misalignment of our exposure definitions with real-world decision-making.

Fourth, time-varying confounders—including concurrent policies, economic disruptions and subsidies, healthcare access changes, trust in government, and social behavioral shifts—co-occurred and likely interacted, making it extremely challenging to disentangle their effects with existing datasets. In response, we deliberately restricted our scope to maximize interpretability, focusing specifically on mental health outcomes following policy removal during “low or declining” pandemic severity periods. Our findings do not address policy restrictiveness, duration, cumulative effects, or lifting during high pandemic severity. Finally, our analyses are restricted to the US which may limit the generalizability of our findings to countries with different sociocultural contexts. Nevertheless, this work may provide insights into the mental health trade-offs of containment policies that could inform preparedness frameworks for future pandemics.

## CONCLUSIONS

Taken together, our findings provide causal evidence that lifting containment policies during low pandemic severity produces immediate but transient mental health benefits. Future pandemic responses may integrate mental health considerations alongside infection control, by accounting for both containment policies and underlying socioeconomic heterogeneity.

## Data Availability

The current study is based on publicly available datasets, namely the Behavioral Risk Factor Surveillance System (BRFSS) and Oxford Covid-19 Government Response Tracker (OxCGRT). Each of these datasets can be accessed via the associated website(https://www.cdc.gov/brfss/annual_data/annual_data.htm) and GitHub repository(https://github.com/OxCGRT/covid-policy-tracker), respectively.

https://github.com/OxCGRT/covid-policy-tracker

## ACKNOWLEDGEMENTS

We thank Drs. Miguel Hernan (Harvard T. H. Chan School of Public Health), Elizabeth Diemer (University of Pennsylvania) for their valuable feedback and guidance throughout the study.

YHL, JWS, SB, AB, JFDLH, DW, YZ, DF, JL, ES, YFL, DF, and COVID-19 Global Mental Health Consortium have been supported by the National Institute of Mental Health (1RF1MH134638).

## CRediT

Conceptualization: YHL, MC, JFDLH, JWS, SB, AB

Data curation: MC, JFDLH

Formal analysis: MC, JFDLH

Funding acquisition: JWS, SB, AB

Investigation: YHL, MC, JFDLH

Methodology: YHL, MC, JFDLH, LDA

Project administration: YHL

Resources: JWS, SB, AB

Software: MC, JFDLH

Supervision: JWS, SB, AB, YHL

Validation: MC, JFDLH

Visualization: MC, JFDLH

Writing – original draft: YHL, MC, JFDLH, JWS

Writing – review & editing: JDT, OVE, GGM, YZ, LDA, PFZ, DF, JL, ES, YFL, DF, DW

