## Supplementary material for "Would Lifting Versus Maintaining COVID-19 Containment Policies Have Reduced Psychological Distress in the US?": Online Supplements

### Online Supplemental Materials

#### eMethods 1: Selecting Containment Policies

Our analysis focuses on three pandemic containment policies: school closures (eFigure 1a), workplace closures (eFigure 1b), and the cancellation of public events (eFigure 1c). These correspond to three of the seven containment policies tracked by OxCGRT.

We excluded restrictions on public gatherings (eFigure 1d), because it was not coded as recommendation versus mandate but instead as restrictions in the permitted size of a group (e.g., 10 people, 100 people) (OxCGRT codebook, 2022). We excluded three additional policies due to their limited adoption throughout the study period, as depicted in eFigures 1e through 1g.

Specifically, closures of public transport (eFigure 1e) were rarely implemented, in part reflecting the limited relevance of statewide transit policy in most US contexts, and in part because public transit was mostly used by essential workers (Hu and Chen, 2021).

Stay-at-home requirements (eFigure 1f) were primarily implemented early in the pandemic, with few states maintaining them beyond the first three months; these orders also varied in regional scope, target populations, and enforcement mechanisms, often relying on individual compliance rather than strict government oversight (Moreland et al., 2020; Bourassa, 2021).

Restrictions on internal movement (eFigure 1g) were also rare, reflecting both constitutional protections on freedom of movement and political reluctance to restrict travel.

**eMethods 2: Estimating Multi-Year Psychological Distress Prevalence**

***Model Specification***

Psychological distress prevalence was estimated by calculating a weighted average of individual-level psychological distress classifications, using the survey weights provided by the BRFSS. As detailed in the Outcomes section in the Main text, psychological distress was defined by responses to question Q2.2 of the BRFSS mental health questionnaire: "*Now thinking about your mental health, which includes stress, depression, and problems with emotions, for how many days during the past 30 days was your mental health not good?*". Individuals who reported 0–14 days of poor mental health were classified as not distressed, while those reporting 15–30 days were classified as distressed.

In accordance with BRFSS guidelines, multi-year prevalence estimates were additionally weighted by the proportion of the total sample contributed by each survey year. This adjustment is expressed as:

$$PsychDis\%_Y = \frac{\sum_y^Y \sum_{i=1}^{N_y} \beta_y w_{y,i} \, PsychDis_{y,i}}{\sum_y^Y \sum_{i=1}^{N} \beta_y w_{y,i}} ; \beta_y = \frac{N_y}{\sum_y^Y N_y}$$

<u>Model Terms:</u>

$PsychDis\%_Y$ - The BRFSS multi-survey year psychological distress prevalence estimate

$Y$- Set of BRFSS survey years

$N_y$- The number of samples that belong to a specific survey year

$\beta_y$ - The proportional adjustment weight for year *y*, based on its sample size contribution to the total multi-year sample

$w_{y,i}$ - BRFSS survey weight for individual *i* in year *y*

$PsychDis_{y,i}$- Dichotomized psychological distress indicator (1 if respondent reported >14 days of poor mental health in past 30 days, 0 otherwise)

#### eMethods 3: Estimating Secular Trends of Psychological Distress

##### *Overview of Psychological Distress Changes*

To estimate changes in psychological distress prevalence at the national and state levels, we compared observed prevalence during the pre-vaccination guidance period with expected prevalence given pre-pandemic trends. For the main analysis, we modeled 7-year pre-pandemic trends (January 2013–December 2019) using weighted linear regression with BRFSS survey weights, then projected this trend to estimate the expected prevalence for the pre-vaccination pandemic period (Figure 2). Sensitivity analyses used 5-year and 9-year trends (9 years being the longest possible span due to changes in BRFSS survey methodology in 2011[39]).

##### *Statistical Tests for Psychological Distress Changes*

P-values and confidence intervals for the national and state-level changes in depression prevalence from April 2020 through April 2021, relative to pre-pandemic trends, were computed as follows. The standard errors for the observed psychological distress prevalence ($SE_{obs}$) and the projected estimate ($SE_{proj}$) were combined to calculate the standard error of the change in psychological distress ($SE_{\Delta PsychDis} = \sqrt{SE_{obs}{}^{2} + SE_{proj}{}^{2}}$). A z-statistic was then computed to estimate 95% confidence intervals, and a z-test was used to calculate the p-value. Because this analysis was conducted across 50 states and 1 national estimate, statistical significance was Bonferroni-corrected as follows: p < .05/51 *, p < .01/51 **, p < .001/51 ***.

##### *Overview of Depression Correlations and Odds Ratio*

In addition, we examined the association between prevalence changes and state characteristics from the 2019 Census using Pearson correlation. We also performed two weighted linear regressions to test the association of daily distress prevalence with (1) daily COVID-19 deaths per 100,000 and (2) full lockdown implementation, controlling for state and month fixed effects.

<u>2019 State-Level Correlations</u>

Confidence intervals and p-values for the 2019 state-level Pearson correlation were estimated using bootstrapping (n=10,000). To account for multiple comparisons across 13 sociodemographic variables, we applied a Bonferroni correction (p < .05/13 *, p < .01/13 **, p < .001/13 ***).

<u>Daily COVID-19 Death Rate and Full Lockdown</u>

We used a weighted linear regression model, using multi-year adjusted BRFSS survey weights, to estimate the impact of daily deaths rate and full lockdown status on psychological distress.

The analysis was based on data from individuals surveyed between April 2020 through April 2021. We fit two separate models, each incorporating a different pandemic-related factor measured on the interview date (*date)* for individual *i* in state *S*. One model included the COVID-19 death rate per 100,000 ($DeathRate_{date,S}$), while the other included an indicator for full lockdown status ($Lockdown_{date,S}$; 1 = fully locked down, 0 = no policy in place, i.e. partial lockdown days were omitted in the analysis ). The equations can be expressed as follows:

Daily Death Rate Model:

$$PsychDis_{date,i} = \beta_{Death} * DeathRate_{day;S} + \alpha_S + \alpha_M + \epsilon$$

Daily Lockdown Model:

$$PsychDis_{date,i} = \beta_{Lock} * Lockdown_{day;S} + \alpha_S + \alpha_M + \epsilon$$

where $PsychDis_{day,i}$ represents psychological distress classification based on the BRFSS mental health questionnaire for individual $i$ on day $date$. $\beta_{Death}$ and $\beta_{Lock}$ are the associated effects between both death rate and full lockdown on psychological distress prevalence respectively. In addition, $\alpha_S$ and $\alpha_M$ denote the fixed effects for all 50 US states and the months in the pandemic respectively, to account for regional and temporal variation. Finally, let $\epsilon$ be the error term.

For the weighted regression, survey weights were adjusted for valid multi-year comparisons by multiplying each weight by the proportion of that year's responses relative to the total number of responses across all years (refer to $\beta_y$ in eMethods 2 for more details). Confidence intervals and p-values are derived from the standard errors of $\beta_{Death}$ and $\beta_{Lock}$provided in the Weighted Linear Model analysis.

#### eMethods 4: Designing Synthetic Control Analysis

##### *Overview*

Using synthetic control (SC) analysis, we estimated the counterfactual trend in psychological distress for a state that lifted full lockdown. Target states refer to those that experienced the policy change, whereas donor states maintained the original policy and were used to construct a counterfactual [40]. Target and donor states were selected through a two-step process to ensure comparability. First, we identified all states that, after imposing full lockdown in March 2020, lifted it only once without reinstatement (28 states). To exclude states that phased out policies over time, we then selected states that had fewer than 100 days of partial lockdown after ending full lockdown (eFigure 3). Five states met all criteria: Maine (166 full lockdown days, 95 additional partial lockdown days), Washington (334 full, 84 partial), New Mexico (354 full, 70 partial), Oregon (370 full, 85 partial), and California (440 full, 45 partial).

Maine served as the target state, lifting full lockdown earliest (September 14, 2020) and during a period of "low/declining" severity, while the other four provided the donor pool for constructing synthetic controls. All donor states maintained full lockdowns for at least five additional months and continued partial lockdowns beyond May 2021. To reduce noise, each monthly estimate used 3-month rolling averages incorporating responses from preceding and following months. Donor weights were derived from data covering the 7 years prior to Maine's full reopening (August 2013 - August 2020). Weights were then applied to estimate Maine's counterfactual for 24 months post-intervention (through September 2022), extending beyond our primary study period (through May 2021) to examine whether target and counterfactual trends converged after initial divergence (Figure 3).

##### *Model Specification*

We estimated counterfactual psychological distress prevalence in Maine using a synthetic control approach, modeling it as a weighted combination of donor states (Washington, New Mexico, Oregon, and California):

$$PsychDis_{ME;m} = w_{WA} PyschDis_{WA;m} + w_{NM} PsychDis_{NM;m} + w_{OR} PsychDis_{OR;m} + w_{CA} PsychDis_{CA;m} + \epsilon$$

where $PsychDis_{S;m}$ is the rolling 3-month average psychological distress prevalence in states at the three month prevalence centered at month $m$ (i.e., averaging months *m – 1, m, and m + 1).* Learned weights assigned to each donor state are denoted as $w_s$. Weights were determined by minimizing the difference between Maine's observed rolling 3-month depression prevalence and that of the synthetic control during the pre-intervention period (August 2013 - August 2020). The trained model was then used to generate counterfactual estimates for the post-intervention period (October 2020 - May 2021).

##### *Sensitivity Analysis*

To evaluate the robustness of our findings, we conducted several sensitivity analyses. First, we performed a leave-one-out analysis by sequentially removing each donor state from the donor pool. We also tested the model using alternative pre-intervention periods, including an extended training window from August 2011 to August 2020 and a shorter window from August 2015 to August 2020.

As shown in eFigure 4, Maine is broadly similar to donor states in most key sociodemographic characteristics in 2019. The main exception is homeownership, where Maine ranks among the highest in the country. Despite this difference, our synthetic control analysis remained highly predictive of the pre-intervention data.

**eMethods 5: Estimating Within-State Fixed Effects of Containment Policies on Psychological Distress**

***Overview***

Across states, we examined the short- and long-term effects of lifting containment policies on psychological distress prevalence using WSFE models, borrowing from case cross-over analysis [41]. As illustrated in Figure 4, we defined periods composed of two separate 30-day windows (pre-treatment and post-treatment), with washout periods of 0, 30, and 60 days between them to test for persistence of the effect. These periods are of two types: a *control period,* where policy is present throughout both windows, and a *case period*, where policy is lifted at the end of the pre-treatment window. Each state may have multiple control and case periods, provided that the pre-treatment window falls during the pre-vaccination guidance period (April 2020 - April 2021) and occurs during a "low/declining" severity period. Additionally, to reduce overlap in survey samples, start dates of different control periods for a given state were required to be at least 7 days apart.

For the models, we regressed the post-treatment prevalence of psychological distress on case/control status, pre-treatment prevalence, and state and month fixed effects. As shown below, models additionally included covariates for prior policy duration and other concurrent policies. To increase statistical power, data points were weighted by the inverse combined variance of the pre- and post-treatment prevalence estimates: $(1 / (SE^2_{pre} + SE^2_{post}))$. We ran this analysis for four containment policies - full lockdown, school closures, workplace closures, and cancellation of public events - resulting in a total of 12 tests. Sensitivity analyses use different pandemic severity classifications as described in the Methods "Pandemic Severity" section.

***Model Specification***

The within-state fixed effects (WSFE) model is as follows:

$$PsychDis\%_{S;Post}^{wash;\,t} = \beta * Treatment_{S;\,pol}^{t} + \gamma * PsychDis\%_{S;Pre}^{t} + \Theta_{t;S} * Covar_S^t + \alpha_S + \alpha_M + \epsilon$$

where $\Theta * Covariates_S^t$ can be further expanded as:

$$\Theta_{t;S} * Covar_S^t = \Theta_0 * PrevPol_{S;Pre;pol}^{t} + \Theta_1 * AltPol_{S;Pre;alt}^{t} + \Theta_2 * AltPol_{S;Post;alt}^{t}$$

<u>Model Terms:</u>

$PsychDis\%_{S;Post}^{wash;\,t}$ –The psychological distress prevalence (percentage) in state *S* during the final 30 days of the experimental period (i.e. post-treatment window), following the treatment assignment at time *t* and a washout period (*wash;* the duration of days between the pre-treatment and post-treatment window).

$Treatment_{S;pol}^{t}$ – A binary indicator denoting whether state *S* at time *t* was assigned treatment (i.e. lifting of policy *pol*) after the first 30 days of the experimental period. Specifically, $Treatment_{S;pol}^{t}$ = 1 when the containment policy was active for the first 30 days of the experimental period but was lifted one day after.

$Treatment_{S;pol}^{t}$ = 0 when the policy remained continuously active across the entire experimental period.

$PsychDis\%_{S;Pre}^{t}$ – The psychological distress prevalence (percentage) in state S during the first 30 days of the experimental period (i.e. pre-treatment window), included to control for baseline differences.

$Covar_S^t$ – Time-varying covariates included the following:

$PrevPol_{S;Pre;pol}^t$ – Days the policy of interest (*pol*) was continuously active before the start of the experimental period for state *S* at time *t*, thereby accounting for the cumulative effect of prolonged exposure to the policy before treatment assignment.

$AltPol_{S;Pre;alt}^t$ – Days an alternative policy (*alt*) was active during the pre-treatment window for state *S* at time *t*.

$AltPol_{S;Post;alt}^t$ – Days an alternative policy (*alt*) was active during the post-treatment window for state *S* at time *t*.

From the four policies examined in the WSFE model - full lockdown, school closures, workplace closures, and canceling of public events - there may be multiple alternative policies depending on the main policy of interest. For example, if the main policy being studied is school closures, then workplace closures and canceling of public events would be considered alternative policies and have both their own $AltPol_{S;Pre;alt}^t$ and $AltPol_{S;Post;alt}^t$ terms. In contrast, when full lockdown is the main policy of interest, there are no alternative policies during the pre-treatment window because, by definition of the study design, all three containment policies (school, workplace, event closures) must be active during that period. During the post-treatment window, however, all policies, but not all three concurrently, could remain active after lifting, thereby requiring three separate $AltPol_{S;Post;alt}^t$ terms.

$\alpha_S$ –State fixed effects controlling for time-invariant factors (e.g., demographics, healthcare access, cultural norms).

$\alpha_M$ – Month fixed effects that account for seasonal patterns in psychological distress and the cumulative impact of living through an extended period of the pandemic.

$\epsilon$ –Error term

$\beta$ –The coefficient of interest, representing the average treatment effect of policy lifting on psychological prevalence outcome, measured in percentage points.

We assigned greater weight to observations with more precise depression change estimates by using the inverse of the squared standard error as weights during model training. To increase statistical power, data points were weighted by inverse combined variance ($1 / (SE^2_{pre} + SE^2_{post})$).

**eMethods 6: Specifying Target Trial Emulation (TTE) Models**

***Overview***

We implemented a target trial emulation to estimate the effects of a sustained containment policy strategy, asking: When pandemic severity is low or declining, what is the effect of lifting versus maintaining a policy on psychological distress over a 90-day period? Unlike dynamic treatment strategies that adapt treatments to follow-up outcomes, we examined a sustained strategy with treatment arms fixed at baseline.

A trial was defined as any period during which a state met the following criteria: (1) occurred during April 2020 - April 2021 (pre-vaccination guidance period) and (2) pandemic severity transitioned to "low/declining" after 30 consecutive days of "high" pandemic severity with active policy (run-in period), (eTable 1). Following eligibility, we tracked policy implementation during a 90-day exposure window. The outcome was assessed as the change in prevalence of psychological distress comparing the 30-day run-in period with the 30-day period following the exposure window. States could contribute multiple observations by initiating new trials at different times, provided they met the eligibility criteria. Trials were censored for the return to "high" severity or policy reinstatement during the exposure window (eFigure 5). Censorship in a trial, however, did not preclude a state from participation in a subsequent trial at a later time. Finally, state-trials were required to have more than 25 responses in both the run-in and follow-up periods to ensure statistical reliability.

We estimated average treatment effects for each policy using parametric G-computation with inverse probability weighting for censoring [42–44]. Models adjusted for 2019 state-level covariates (education and homeownership), days since the pandemic began (March 13, 2020), and a binary indicator for whether the policy remained beyond 14 days in the follow-up window. We were unable to adjust for seasonality due to positivity issues, though it remains a potentially important confounder.

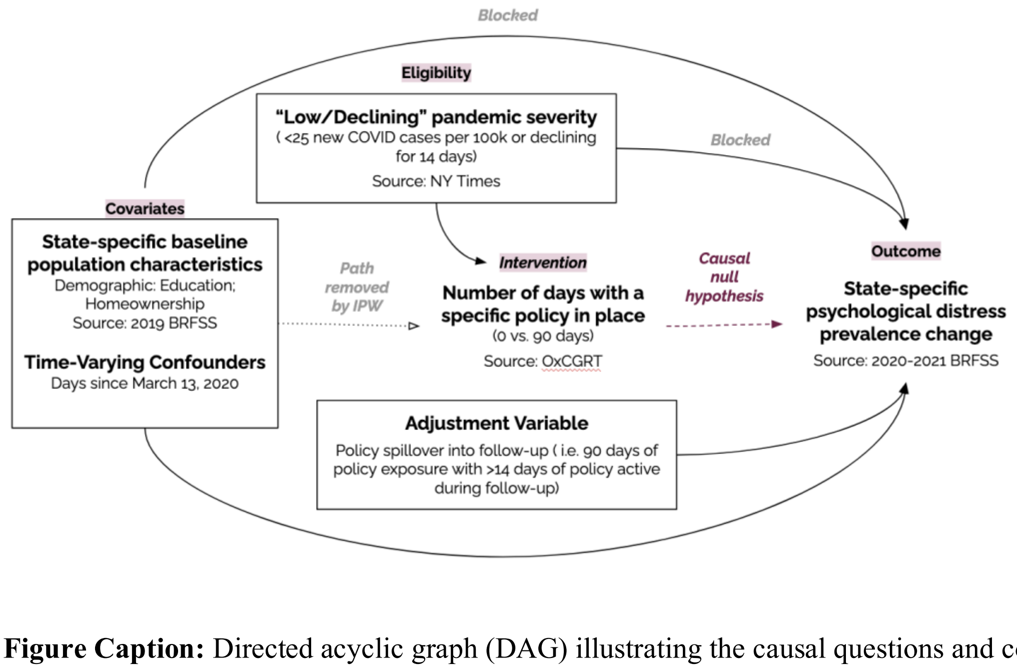


**Figure Caption:** Directed acyclic graph (DAG) illustrating the causal questions and covariates being accounted for in the TTE analysis. Boxes around covariates indicate regression adjustment, a dotted line indicates a path removed using inverse probability weighting, and a dashed line indicates causal null hypothesis being evaluated.

##### *Model Specification*

Based on the causal diagram shown above, the TTE model is fitted as follows:

$$\Delta PsychDis\%_S^{Out;\,t} = \alpha + \beta * PolicyDays_{S;pol}^{Exp;\,t} + \lambda * StillOn_{pol} + \Theta_S * Covar_S^{2019} + \Theta_t * Covar_t$$

while the IP weighting was fitted as follows:

$$IsCensored_S^t = logistic(\alpha + \Phi_S * Covariates_S^{2019} + \Phi_t * Covariates_t\ )$$

Model Terms:

$\Delta PsychDis\%_S^{Out;\,t}$ – percentage point change in psychological distress prevalence between the 30-day run-in period ($PsychDis\%_S^{RunIn;\,t}$) and the 30-day follow-up period ($PsychDis\%_S^{FollowUp;\,t}$) for state $S$ at time *t*, calculated as $PsychDis\%_S^{FollowUp;\,t} - PsychDis\%_S^{RunIn;\,t}$.

$PolicyDays_{S;\,pol}^{Exp;\,t}$ – The primary exposure variable representing the number of days (0-90) that a containment policy *pol* (i.e. full lockdown, school closure, workplace closure, and canceling of public events) remained in place under low pandemic severity conditions during the exposure window for state *S* at time *t*. This is the "dose" of policy exposure, where higher values indicate longer sustained policy implementation (during "low or declining" severity periods).

$StillOn_{pol}$ – A binary indicator for whether the policy of interest remained active for more than 14 days during the follow-up window. This accounts for the possibility that the policy was not fully lifted and may still have influenced outcomes.

$Covar_S$ – A vector of 2019 state-level covariates was sourced from the US Census Bureau. For our analysis, we adjusted for two covariates: the percentage of the population with a bachelor's degree or higher, and the proportion of home ownership in state *S*. These were the only covariates found to be significantly associated with changes in psychological distress, based on a correlation analysis (Figure 2c) between 2019 state-level characteristics and the change in psychological distress from April 2020 through April 2021.

$Covar_t$ – A vector of time-varying covariates. Due to sample size limitations, we included only one: the number of days since the beginning of the pandemic (May 13, 2020), to account for the passage of time over the course of the pandemic.

$\beta$ – The coefficient of interest, representing the average effect of 90-days of sustained policy implementation on depression prevalence change.

$IsCensored_S^t$ – A binary indicator for whether a state-trial was censored. Censorship was defined as occurring when either: (1) the pandemic severity in the state shifted to "high" during the exposure window, or (2) the policy of interest was reintroduced after having been lifted. The output of the model for all state-trials was then used to create the inverse probability weights for the main linear regression in the TTE analysis

##### *Sensitivity Analysis*

We performed several sensitivity analyses. First, we tested alternative pandemic severity classifications, using thresholds of 10 and 50 cases per 100,000 to define "high" and "low/declining" severity. We also used different methods to account for policy spill-over into the follow-up window, i.e. states with policies active for 90 days in the exposure window and >14 days in the follow up window. Specifically, sensitivity tests excluded state-trials with >14 days of spillover or removed the indicator as part of the model entirely. We further assessed the impact of including 2019 state-level poverty rates, which showed a strong association with psychological distress in the first year of the pandemic albeit not a significant one. Lastly, we experimented with a 30-day washout period to evaluate the effect of greater temporal separation between the exposure and follow-up window.

##### *Power Analysis*

A power analysis was conducted to assess the statistical power of the TTE (eFigure 12). Sample sizes ranging from 10 to 100 were evaluated, with draws designed to mimic the sample size distribution observed in the primary analyses (full lockdown: $n = 18$; school closure: $n = 59$; workplace closure: $n = 29$; cancellation of public events: $n = 25$). Effect sizes were specified in terms of Cohen's $d$ and subsequently transformed into changes in the prevalence of psychological distress. This transformation was performed by multiplying Cohen's $d$ by the standard deviation of the outcome. To ensure a conservative and consistent scaling across analyses, standardized effect sizes were converted into percentage-point changes using 4.7 percentage points, the largest observed outcome standard deviation (full lockdown: 4.26%, school closure: 4.03%, workplace closure: 4.70%, cancellation of public events: 4.05%).

**eFigure 1: Timeline of COVID-19 Containment Policy Implementation across US States.**

Each panel shows the implementation of a containment policy across all 50 US states from March 2020 to May 2022. Horizontal lines indicate periods when each state had the respective policy in effect as a requirement (OxCGRT level ≥2), rather than a recommendation. Blue star-shaped markers indicate the date each state reached 25% COVID-19 vaccination coverage. The vertical dashed line marks the date the CDC issued guidance for fully vaccinated individuals. The light orange background marks the timeframe eligible for analysis, spanning the period from April 2020 to April 2021.

The policies are (a) school closures, (b) workplace closures, (c) cancellation of public events, (d) restrictions on public gatherings, (e) public transport closures, (f) stay-at-home requirements, (g) restrictions on internal movement, (h) full lockdown (i.e. simultaneous school, workplace, and public event closure).

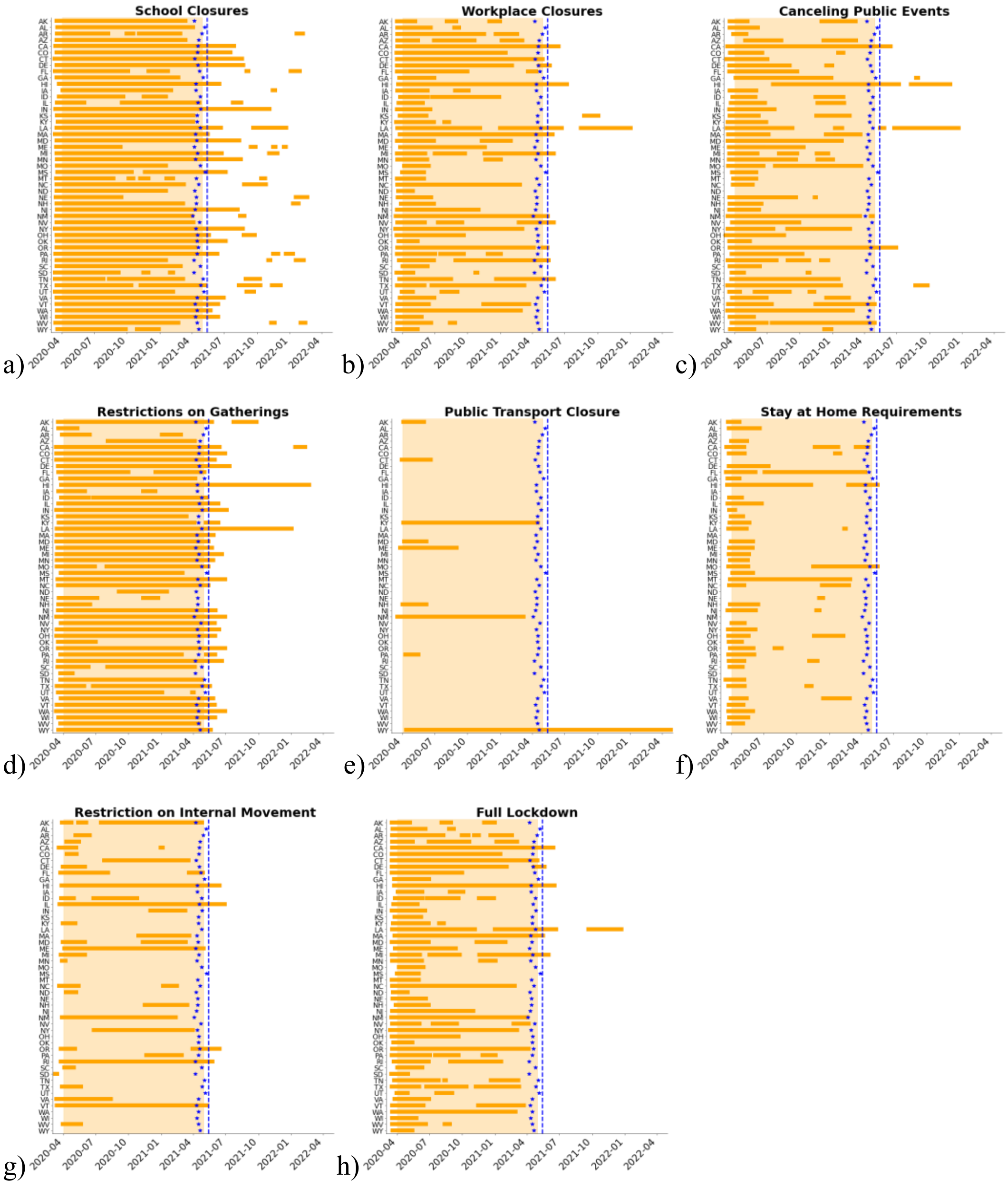

**eFigure 2: Defining Pandemic Severity throughout the COVID-19 Pandemic.**
**(a)** COVID-19 case rates per 100,000 in Washington from March 13, 2020 to May 12, 2021. Shaded red regions indicate periods classified as "high" pandemic severity, as defined in the Methods ("Pandemic Severity" section). A period of "low or declining" severity begins when case rates fall below 25 per 100,000 or show 14 consecutive days of decline. This period ends if a single-day spike exceeds 1.33× the previous day's rate or if a week-long increase in cases above 25 per 100,000, triggering a "high" severity period. *Bottom:* Detected "high" severity days using different algorithm parameters. The default denotes the definition used in the main analysis.

**(b)** Average correlation of "high" pandemic day classifications across US states under different severity definitions. Lower correlations occurred with changes to the minimum case rate threshold, indicating greater sensitivity to this parameter.

**(c - e)** Comparison of "high" severity days under minimum case rate thresholds of 10, 25 (default), and 50 cases per 100,000. The orange shaded area marks the exposure window (April 2020 to April 2021), ending just before the CDC's May 13, 2021 guidance for fully vaccinated individuals (dashed blue line).

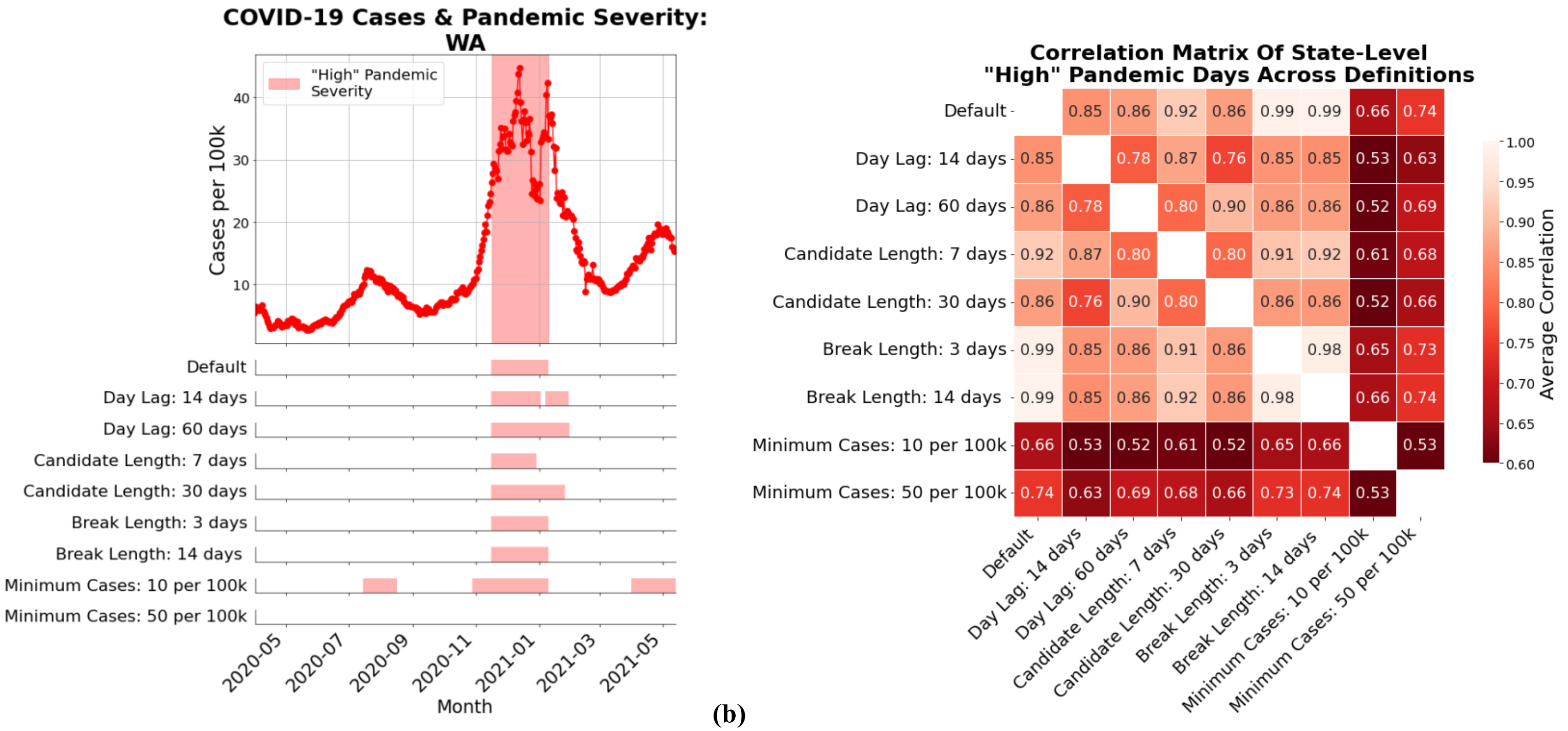

COVID-19 Cases & Pandemic Severity: WA
"High" Pandemic Severity
Cases per 100k
10
20
30
40
Default
Day Lag: 14 days
Day Lag: 60 days
Candidate Length: 7 days
Candidate Length: 30 days
Break Length: 3 days
Break Length: 14 days
Minimum Cases: 10 per 100k
Minimum Cases: 50 per 100k
2020-05
2020-07
2020-09
2020-11
2021-01
2021-03
2021-05
Month
Correlation Matrix Of State-Level "High" Pandemic Days Across Definitions
Default 0.85 0.86 0.92 0.86 0.99 0.99 0.66 0.74
Day Lag: 14 days 0.85 0.78 0.87 0.76 0.85 0.85 0.53 0.63
Day Lag: 60 days 0.86 0.78 0.80 0.90 0.86 0.86 0.52 0.69
Candidate Length: 7 days 0.92 0.87 0.80 0.80 0.91 0.92 0.61 0.68
Candidate Length: 30 days 0.86 0.76 0.90 0.80 0.86 0.86 0.52 0.66
Break Length: 3 days 0.99 0.85 0.86 0.91 0.86 0.98 0.65 0.73
Break Length: 14 days 0.99 0.85 0.86 0.92 0.86 0.98 0.66 0.74
Minimum Cases: 10 per 100k 0.66 0.53 0.52 0.61 0.52 0.65 0.66 0.53
Minimum Cases: 50 per 100k 0.74 0.63 0.69 0.68 0.66 0.73 0.74 0.53
Average Correlation
1.00
0.95
0.90
0.85
0.80
0.75
0.70
0.65
0.60

**(a)** **(b)**

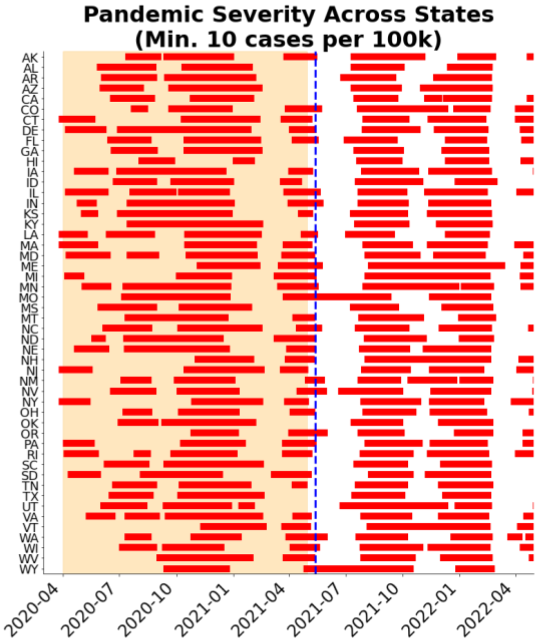
Pandemic Severity Across States
(Min. 10 cases per 100k)
AK AL AR AZ CA CO CT DE FL GA HI IA ID IL IN KS KY LA MA MD ME MI MN MO MS MT NC ND NE NH NJ NM NV NY OH OK OR PA RI SC SD TN TX UT VA VT WA WI WV WY
2020-04 2020-07 2020-10 2021-01 2021-04 2021-07 2021-10 2022-01 2022-04
(c)

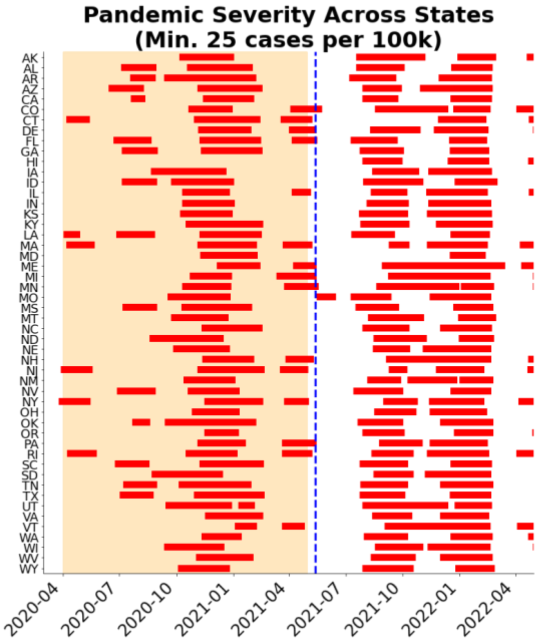
Pandemic Severity Across States
(Min. 25 cases per 100k)
AK AL AR AZ CA CO CT DE FL GA HI IA ID IL IN KS KY LA MA MD ME MI MN MO MS MT NC ND NE NH NJ NM NV NY OH OK OR PA RI SC SD TN TX UT VA VT WA WI WV WY
2020-04 2020-07 2020-10 2021-01 2021-04 2021-07 2021-10 2022-01 2022-04
(d)

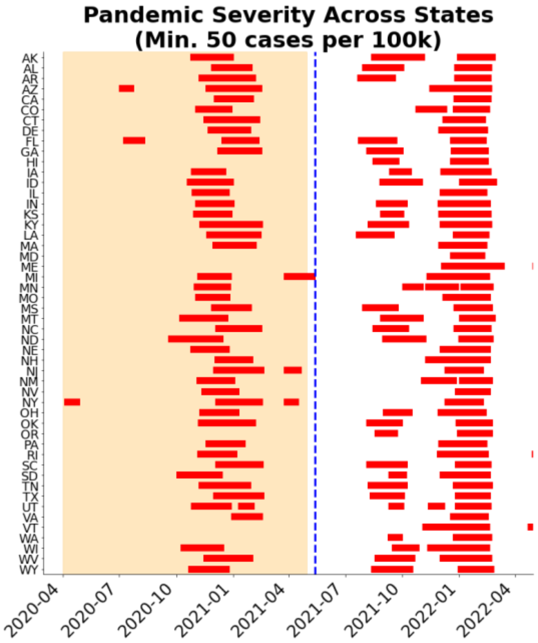
Pandemic Severity Across States
(Min. 50 cases per 100k)
AK AL AR AZ CA CO CT DE FL GA HI IA ID IL IN KS KY LA MA MD ME MI MN MO MS MT NC ND NE NH NJ NM NV NY OH OK OR PA RI SC SD TN TX UT VA VT WA WI WV WY
2020-04 2020-07 2020-10 2021-01 2021-04 2021-07 2021-10 2022-01 2022-04
(e)

**eFigure 3: Selection Threshold for Partial Lockdown Duration in Synthetic Control Analyses.** Distribution of partial lockdown days following the lifting of full lockdown across all eligible states used in the synthetic control analysis. The red dashed line marks the 100-day threshold, which was used as the cutoff for inclusion in the analysis.

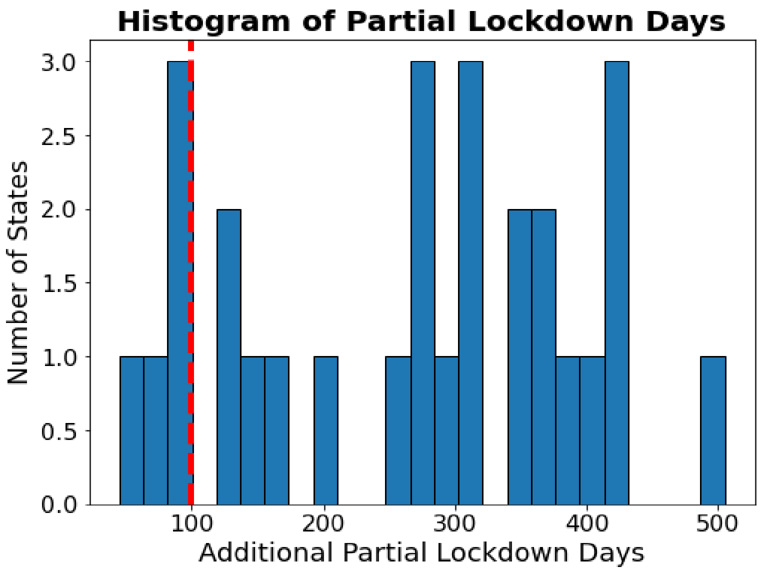

**eFigure 4: Comparison of Target and Donor States on Key Sociodemographic Characteristics Used in Synthetic Control Analyses.** Distribution of 2019 state-level socio-demographic covariates that were significantly associated with psychological distress outcomes when compared to all baseline trajectories (i.e. 5-year, 7-year, and 9-year pre-pandemic trends). Variables shown include: % of the population with a bachelor's degree or higher, % of homeowners, and % below the poverty line. The target state, Maine, is shown in solid red, and donor states (including Washington, New Mexico, Oregon, and California) used in the synthetic control analysis are shown in dashed blue.

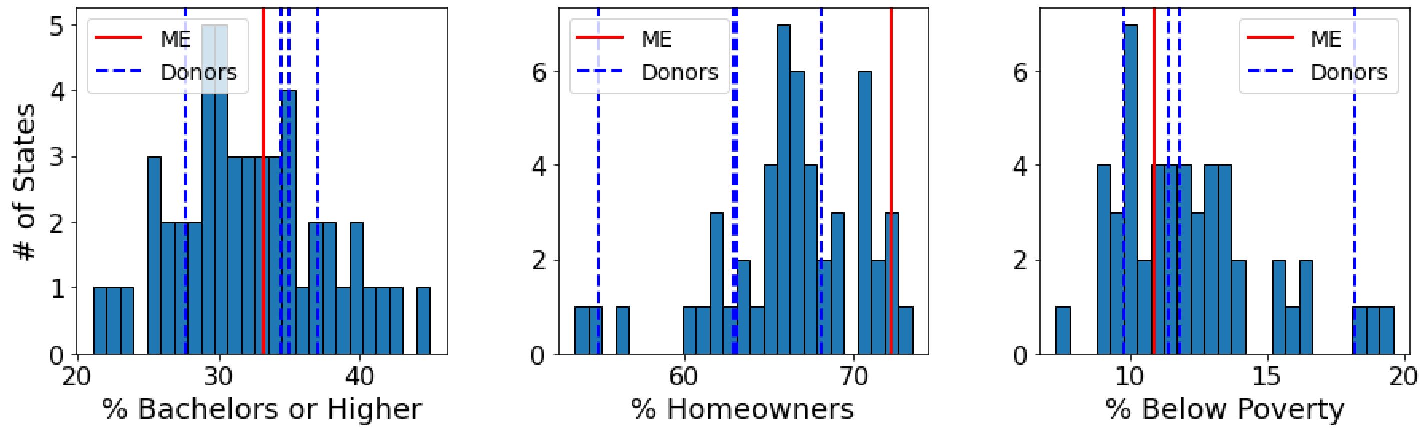

**eFigure 5: Study Designs for Target Trial Emulation Analyses.**
**(a) Hypothetical state examples:** States may experience different durations of policy exposure during the 90-day intervention window. For example, State A lifts policies early ("minimal exposure"), State B lifts policies midway ("moderate exposure"), and State C maintains policies throughout ("sustained exposure").

**(b) Eligibility and censoring scenarios:** Hypothetical examples of states that were either ineligible to enter the trial, censored due to a return to high pandemic severity or policy changes during the exposure window, or completed the trial without interruption.

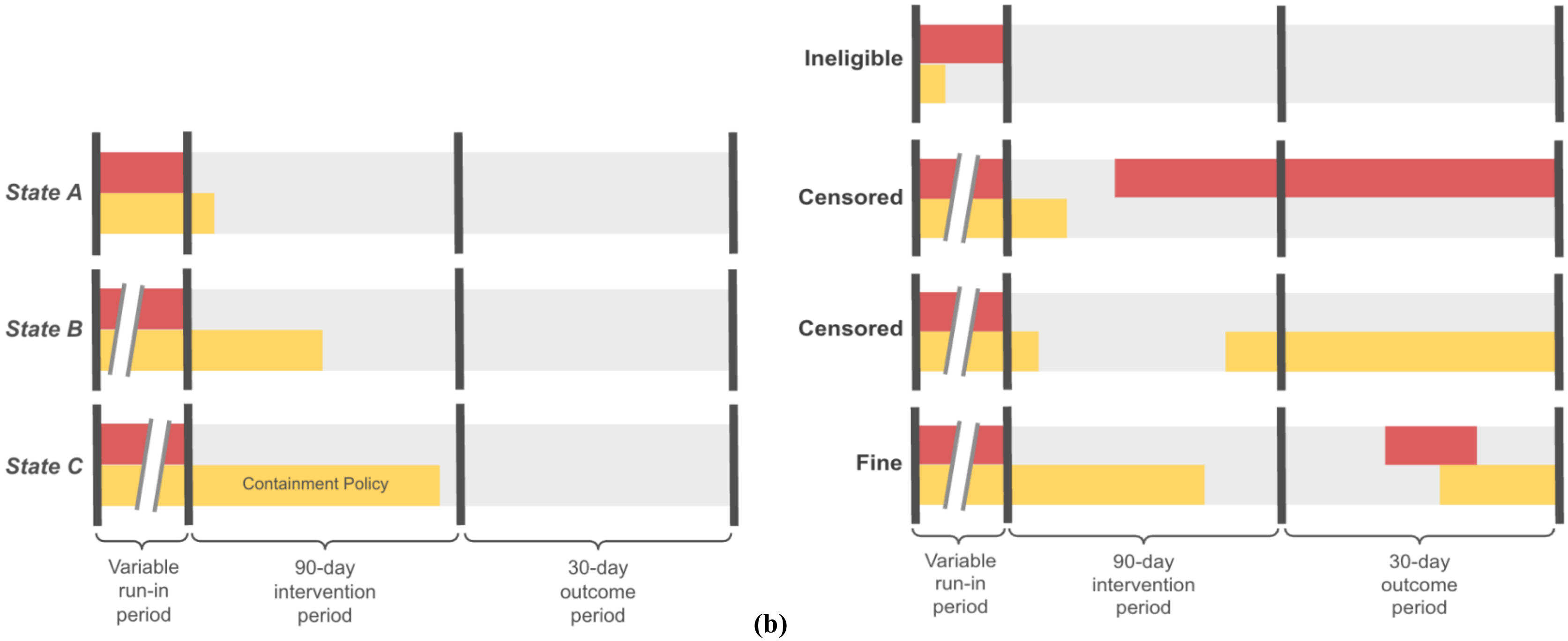


(a) (b)

**eFigure 6: Estimating Sensitivity of Psychological Distress Changes to Baseline Trend Length and Regional Variation.**

**(a - b)** Total change in psychological distress prevalence during the pre-vaccination guidance period (April 2020 to April 2021), calculated as the difference between observed and projected values using **(a)** a 5-year and **(b)** a 9-year baseline, in each US state (including the overall US, indicated as "USA"). Statistical significance was assessed using Bonferroni-corrected thresholds: $p < 0.05/51$ *, $p < 0.01/51$ **, $p < 0.001/51$ **.

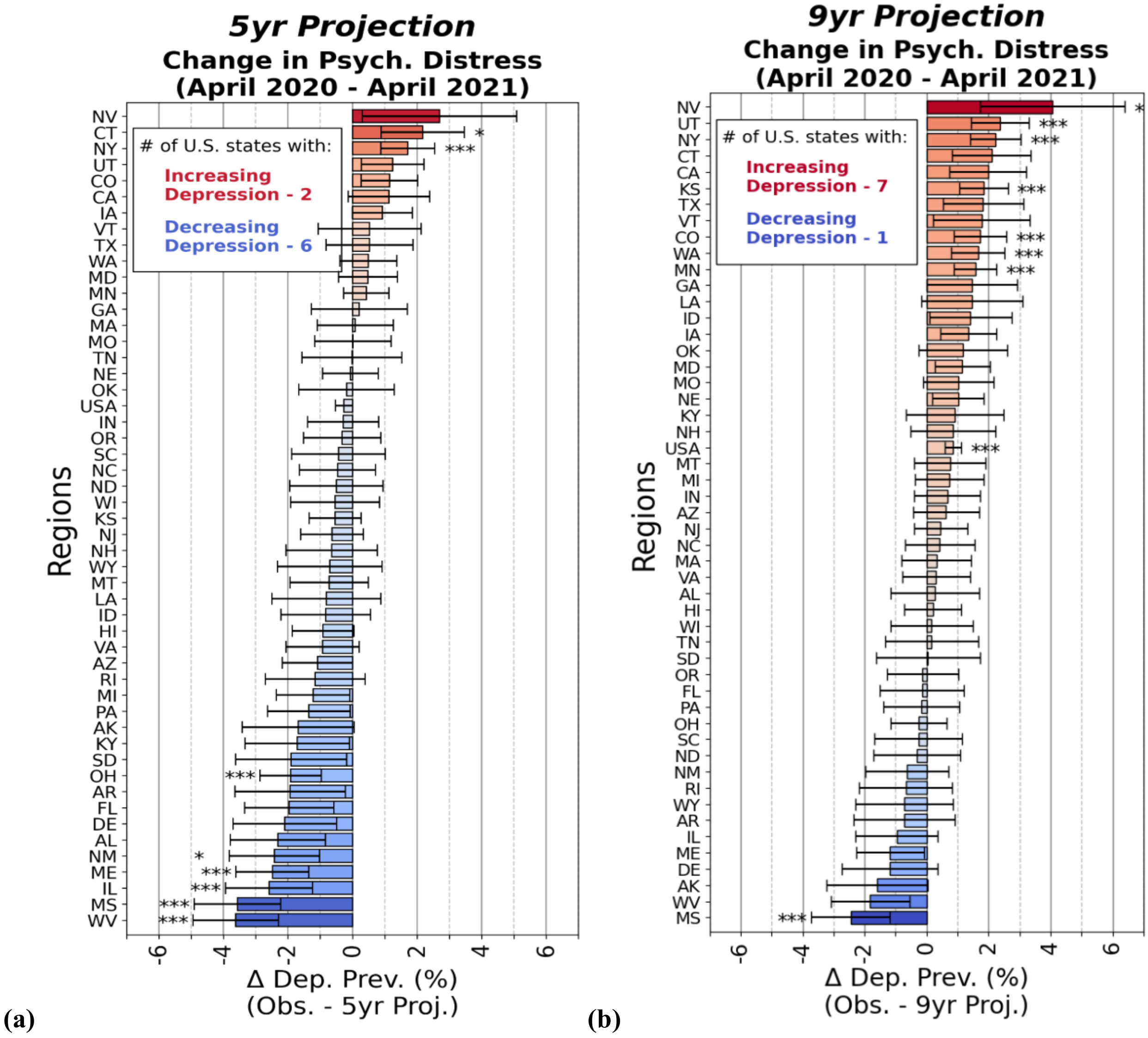

**(c - d)** Correlation between 2019 state-level psychological distress and the change in distress during the pre-vaccination guidance period, using **(c)** a 5-year baseline trend and **(d)** a 7-year trend. Statistical significance was assessed using Bonferroni-corrected thresholds: $p < 0.05/12$ *, $p < 0.01/12$ **.

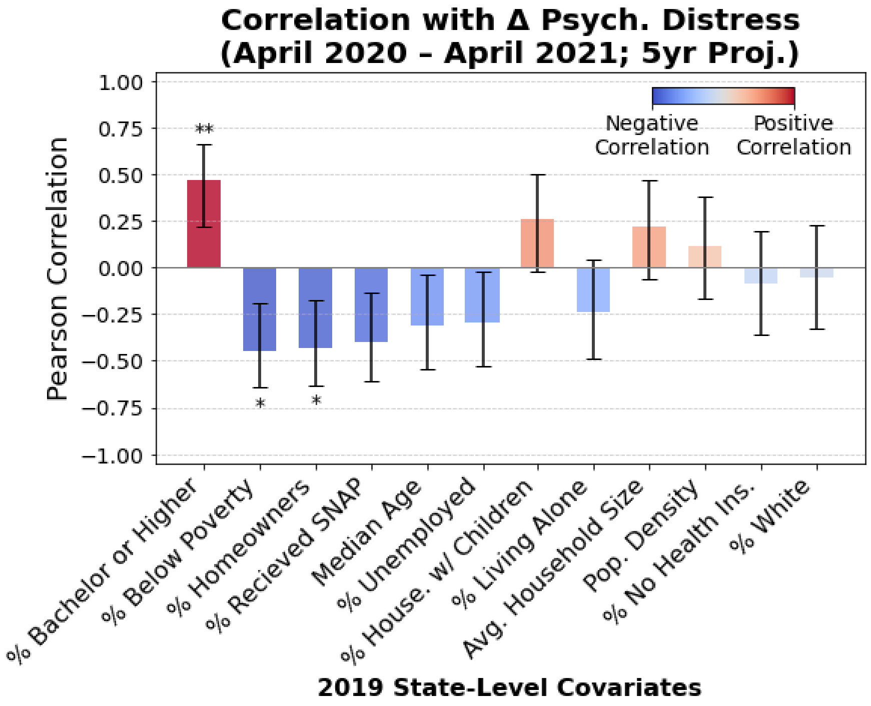


**(c)**

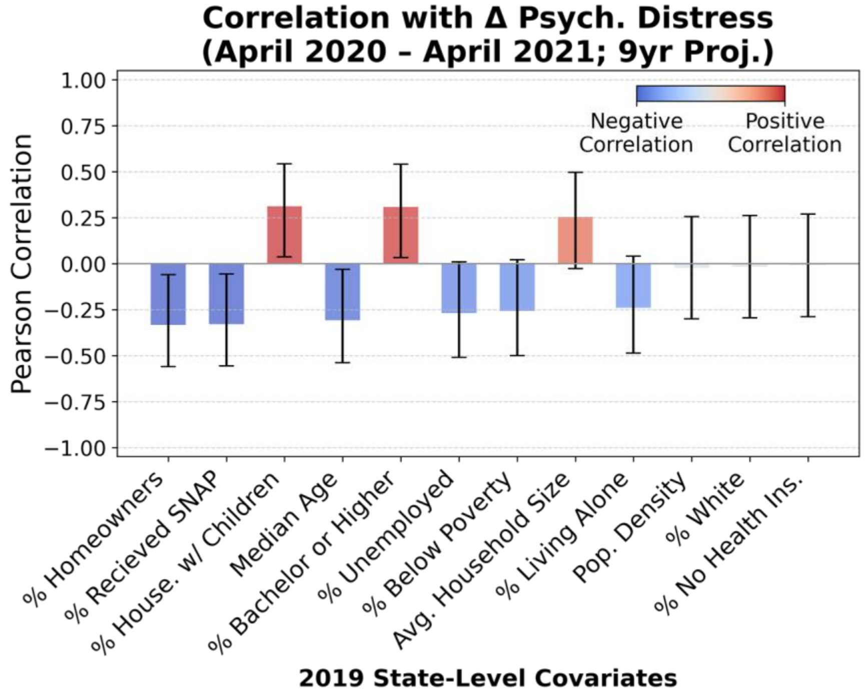


**(d)**

**eFigure 7: Pandemic-Related Dynamics and Psychological Distress in Hawaii.**
**(a)** Monthly psychological distress (top), COVID-19 cases per 100,000 (upper middle), vaccination rate (lower middle), and active containment policies (bottom) in Hawaii from January 2020 to December 2021.

**(b)** Distribution of COVID-19 deaths per 100,000 (top), cases per 100,000 (upper middle), number of "high" severity pandemic days (lower middle), and number of full lockdown days across all 50 US states from April 2020 to April 2021 (bottom). Horizontal lines indicate Hawaii's values.

**(c)** Monthly psychological distress in Hawaii from January 2013 to December 2023, along with the projected 7-year distress (dashed black line, 95% CI in shaded gray) trajectory based on data from January 2013 to 2019.

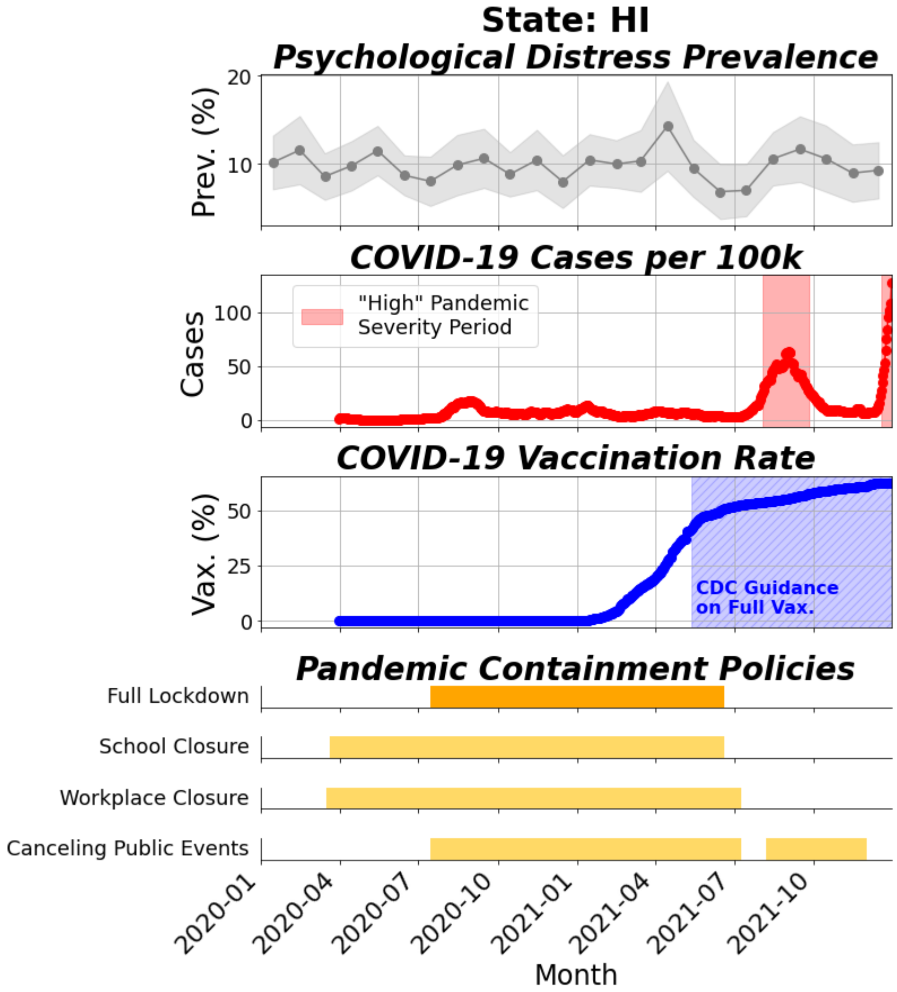


(a)

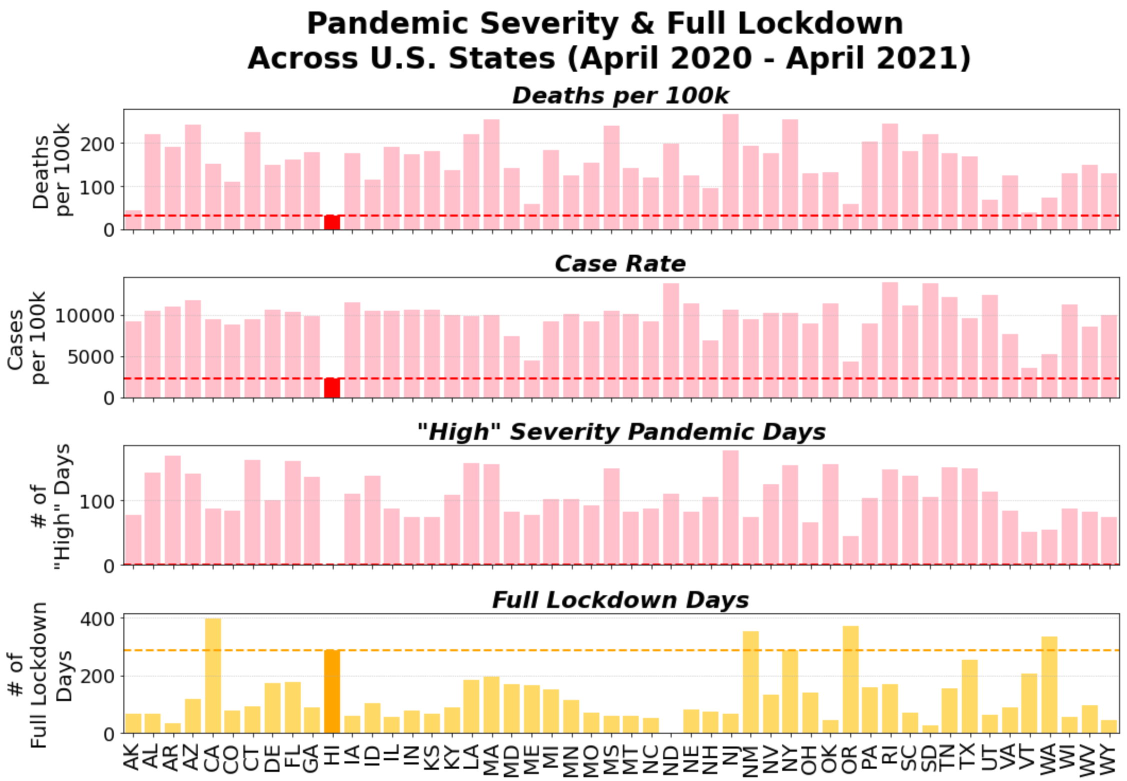
Pandemic Severity & Full Lockdown
Across U.S. States (April 2020 - April 2021)
Deaths per 100k
Deaths per 100k
Case Rate
Cases per 100k
"High" Severity Pandemic Days
### of "High" Days
Full Lockdown Days
### of Full Lockdown Days
AK AL AR AZ CA CO CT DE FL GA HI IA ID IL IN KS KY LA MA MD ME MI MN MO MS MT NC ND NE NH NJ NM NV NY OH OK OR PA RI SC SD TN TX UT VA VT WA WI WV WY

(b)

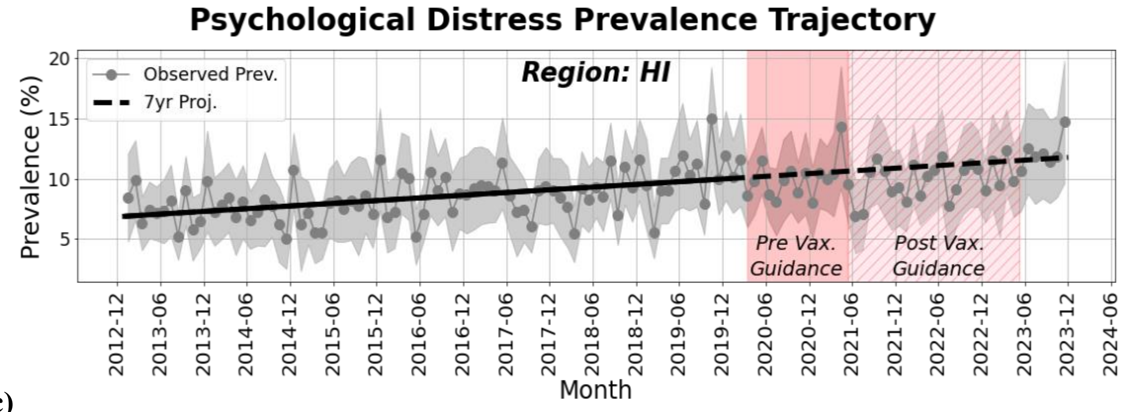
Psychological Distress Prevalence Trajectory
Observed Prev.
7yr Proj.
Region: HI
Pre Vax. Guidance
Post Vax. Guidance
Prevalence (%)
Month

(c)

**eFigure 8: Sensitivity Analysis of Synthetic Control Estimates for Maine.**
Synthetic control analysis of 3-month rolling psychological distress prevalence in Maine, including 95% confidence intervals.

Maine is used as the target state, with Washington, New Mexico, Oregon, and California as donor states. Three different training windows are evaluated: a 5-year window (September 2015 – October 2020, top), a 7-year window (September 2013 – October 2020, middle, used as the main analysis), and a 9-year window (September 2011 – October 2020, bottom). The full training and post-intervention testing period through May 2023 are shown.

The dashed black line represents the primary synthetic control estimate (counterfactual), while the gray dashed lines represent alternative counterfactuals generated through leave-one-out sensitivity analyses. The blue vertical line marks the date Maine lifted its full lockdown (September 14, 2020).

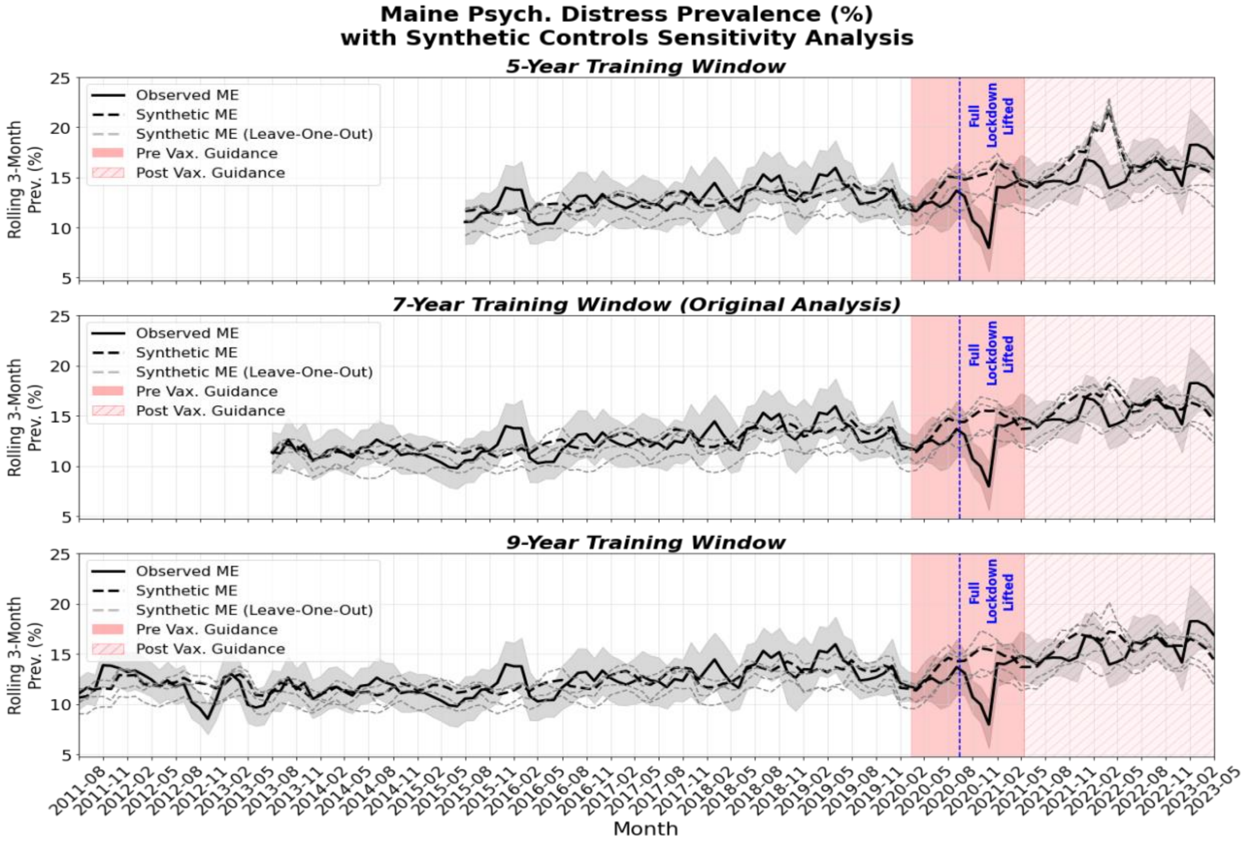

**eFigure 9: Sensitivity Analysis of Within-State Fixed Effects to Pandemic Severity Definitions.** Sensitivity analysis of the within-state fixed effects results (described in the "Within-State Fixed Effects Analysis" section), using two alternative thresholds for defining "high" pandemic severity: **(a)** minimum of 10 cases per 100,000 and **(b)** minimum of 50 cases per 100,000. These definitions affect which time periods are eligible for analysis, as eligibility requires the first 30 days of a period with no occurrences of "high" pandemic severity.

***"High" Pandemic Severity: Min. 10 Cases per 100k***

**Within-State Fixed-Effects**

| Variable | ΔDep.(%) | N | N Cases | P-val |
|---|---|---|---|---|
| **0 to 30 Days After Lifting Policy** | | | | |
| Full Lockdown | -5.26 | 227 | 30 | 0.008* |
| School Closure | 0.05 | 753 | 13 | 0.933 |
| Workplace Closure | -0.21 | 403 | 22 | 0.719 |
| Canceling Public Events | -1.59 | 300 | 32 | 0.006* |
| **30 to 60 Days After Lifting Policy** | | | | |
| Full Lockdown | -2.94 | 161 | 25 | 0.195 |
| School Closure | 0.67 | 695 | 12 | 0.329 |
| Workplace Closure | 0.81 | 325 | 18 | 0.433 |
| Canceling Public Events | -2.46 | 233 | 27 | 0.031 |
| **60 to 90 Days After Lifting Policy** | | | | |
| Full Lockdown | -1.28 | 136 | 24 | 0.437 |
| School Closure | 0.21 | 628 | 12 | 0.769 |
| Workplace Closure | -0.69 | 272 | 14 | 0.557 |
| Canceling Public Events | -2.71 | 200 | 27 | 0.032 |

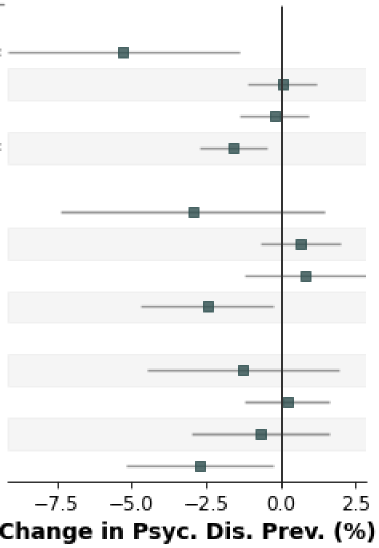


**(a)**

***"High" Pandemic Severity: Min. 50 Cases per 100k***

**Within-State Fixed-Effects**

| Variable | ΔDep.(%) | N | N Cases | P-val |
|---|---|---|---|---|
| **0 to 30 Days After Lifting Policy** | | | | |
| Full Lockdown | -5.69 | 439 | 49 | 0.000** |
| School Closure | -0.85 | 1594 | 23 | 0.049 |
| Workplace Closure | -0.28 | 769 | 42 | 0.477 |
| Canceling Public Events | -1.03 | 665 | 47 | 0.004* |
| **30 to 60 Days After Lifting Policy** | | | | |
| Full Lockdown | -3.71 | 303 | 43 | 0.011* |
| School Closure | -0.18 | 1483 | 17 | 0.739 |
| Workplace Closure | -0.16 | 615 | 37 | 0.815 |
| Canceling Public Events | -0.62 | 517 | 39 | 0.276 |
| **60 to 90 Days After Lifting Policy** | | | | |
| Full Lockdown | -0.62 | 246 | 42 | 0.655 |
| School Closure | -0.48 | 1362 | 16 | 0.373 |
| Workplace Closure | -0.11 | 522 | 30 | 0.908 |
| Canceling Public Events | -1.49 | 419 | 38 | 0.065 |

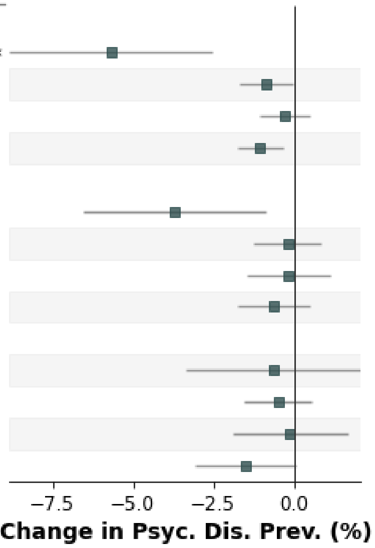


**(b)**

**eFigure 10: Examples of Real Trial Periods for Full Lockdown Containment Policies.** Observed exposure and outcome periods for states participating in trials related to full lockdown (i.e. simultaneous closure of schools, workplace, and public events) illustrating the duration of policy exposure during periods of low pandemic severity. States could contribute multiple trial entries at different time points. Asterisks (*) denote state-trials that were censored either due to a return to a "high" pandemic severity period or because policies were reinstated after being lifted.

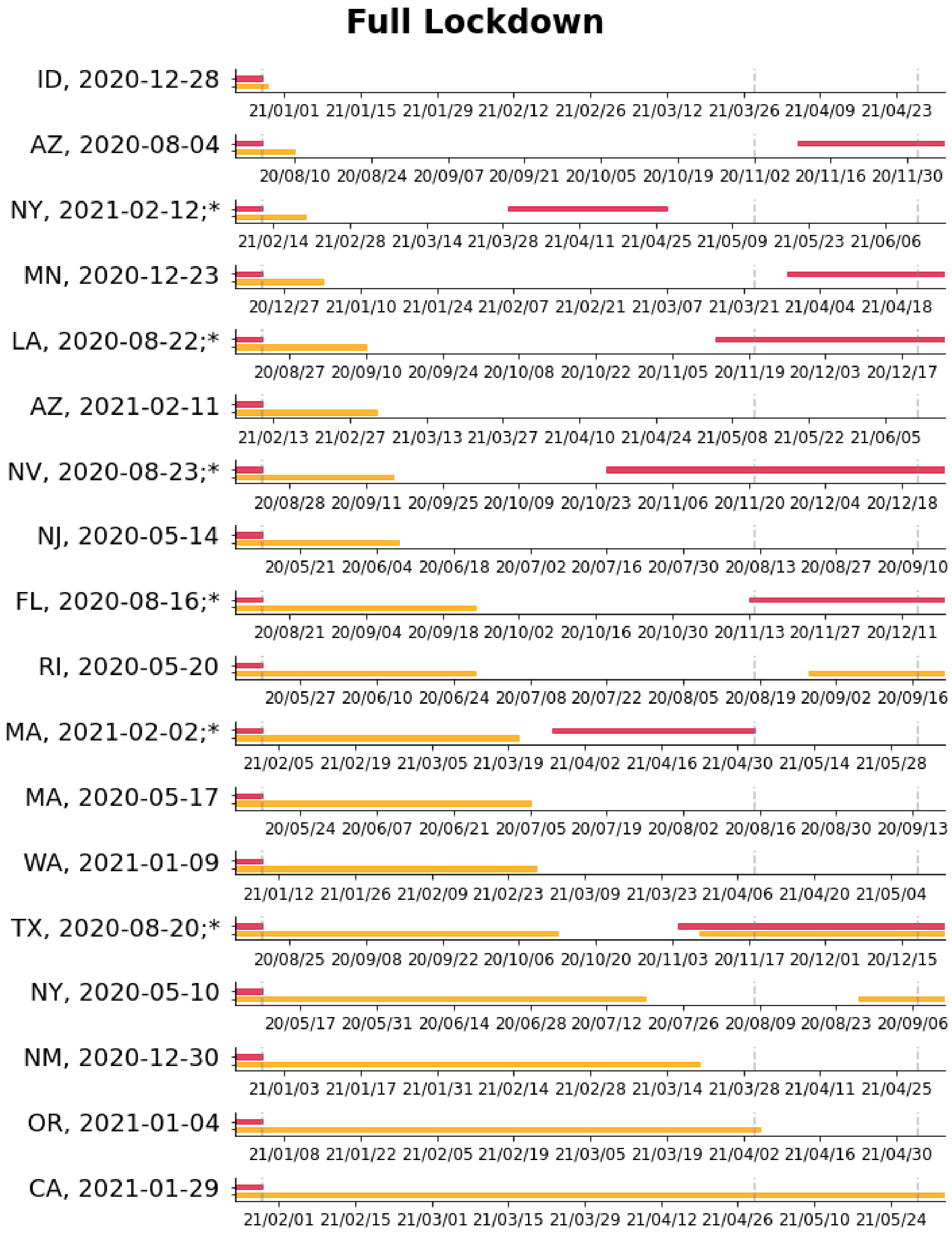

**eFigure 11: Timing of Containment Policy Lifting During Low-Severity Pandemic Periods.** Distribution of the number of days each containment policy was maintained after a state transitioned from a “high” to a “low/declining” pandemic severity period, as part of the target trial emulation. Policies analyzed include full lockdowns, school closures, workplace closures, and cancellation of public events. Light-blue bars represent censored state-trials, while blue bars indicate the non-censored state-trials. Censorship is defined as either (1) a return to high pandemic severity during the exposure window, or (2) the reimplementation of a policy that had previously been lifted during the exposure window.

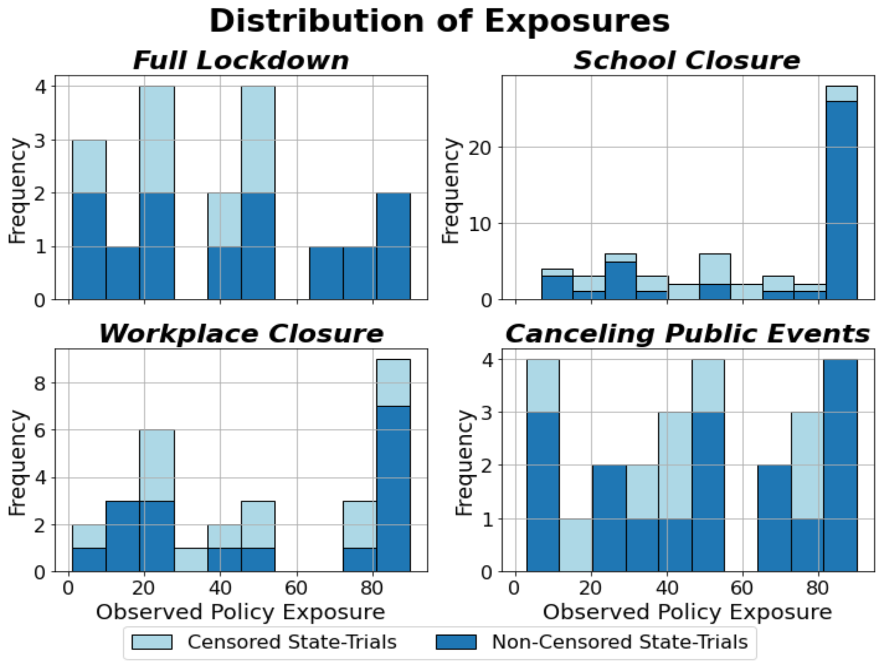

**eFigure 12: Power Analyses for the Target Trial Emulation (TTE)**
A figure showing the statistical power of the TTE across varying sample sizes and effect sizes. We express effect sizes in terms of percentage-point change as described in eMethods 6.

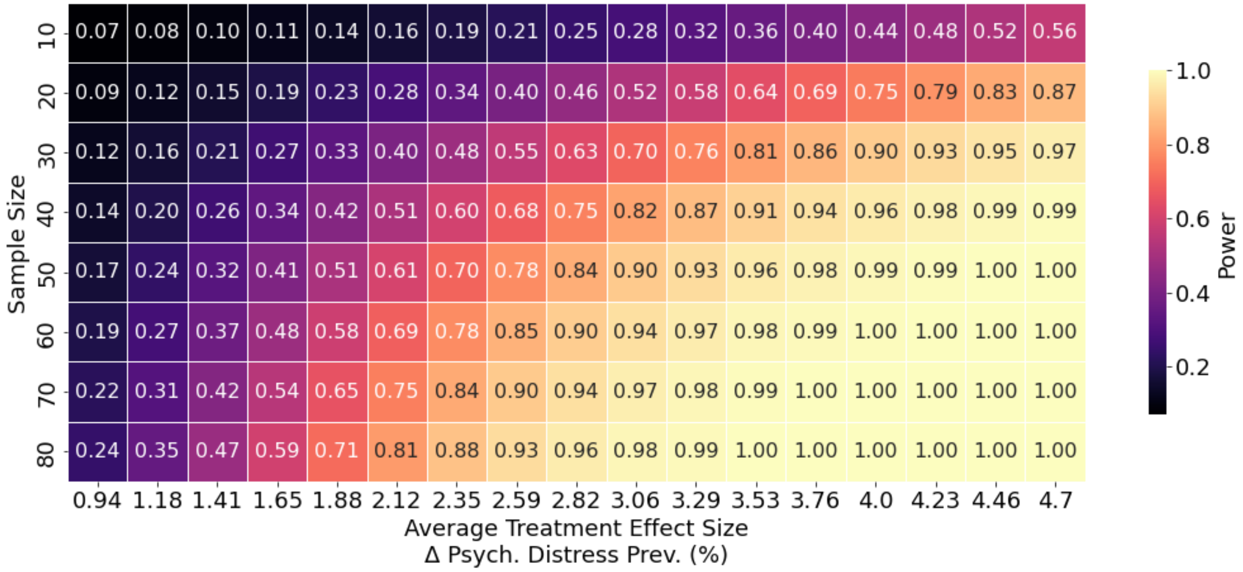

**eFigure 13: Sensitivity Analyses for Target Trial Emulation (TTE)**

This figure presents sensitivity analyses for the TTE outcome. Panels a and b vary the definition of "high" pandemic severity used for trial eligibility: (**a)** a minimum incidence of 10 cases per 100,000 and (**b)** a minimum of 50 cases per 100,000. Panels (**c)** and (**d)** assess alternative strategies for handling policy exposure that extends into the follow-up period: **c)** excluding the binary indicator for policies maintained for the full 90-day exposure and extending more than 14 days into follow-up, and (**d)** restricting the sample to state-trials in which the policy ended within two weeks after the start of the follow-up period. Panel **(e)** adjusts for state-level poverty, a covariate identified as nominally significant in prior analyses. Panel (**f)** evaluates temporal sensitivity by introducing a 30-day washout period between the end of the exposure period and the start of follow-up.

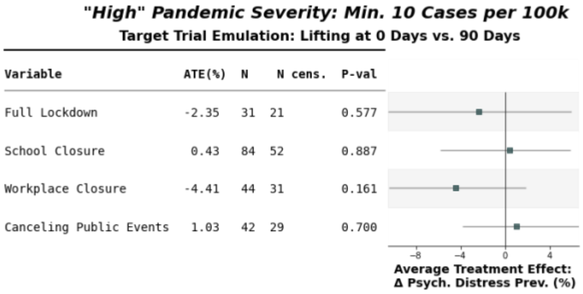


(a)

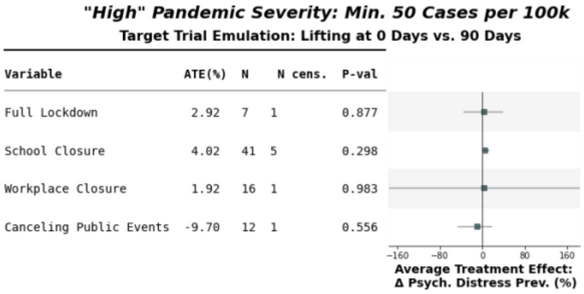


(b)

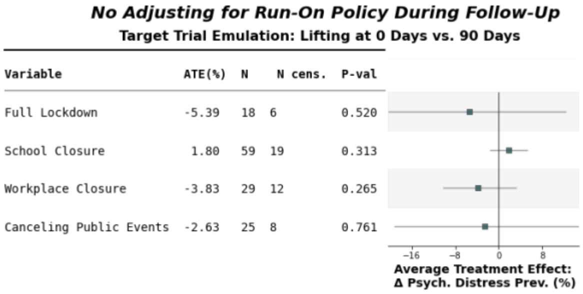


(c)

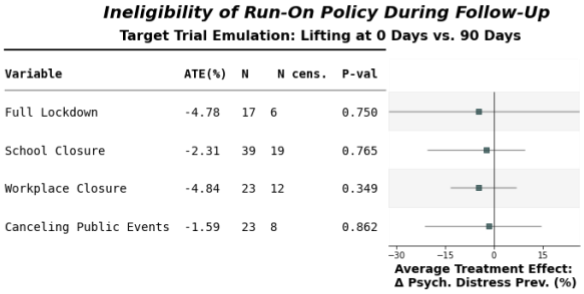


(d)

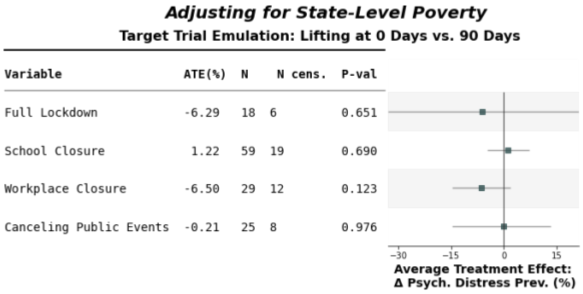


(e)

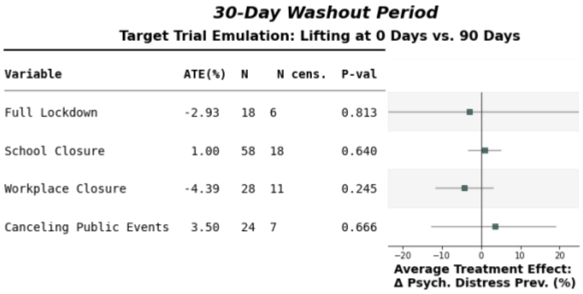


(f)

**eTable 1: Specification and Emulation of a Target Trial for Each Policy Based on Study Data.**

| | Target Trial Specification | Emulation |
|---|---|---|
| Eligibility Criteria | *Inclusion:* US states (n=50) with<br>- 30 consecutive days of high pandemic severity (30 day run-in period, cycle phase 1 Figure 1c)<br>- Active containment policies during the high severity 30-day run-in period<br>- No guidance for fully vaccinated individuals<br>*Exclusion:* States lifting containment policies before transitioning from "high" to "low/declining" severity period | Applied criteria using:<br>- NYTCDR case data<br>- OxCGRT policies<br>- Before May 13, 2021 (Date of CDC guidance for fully vaccinated individuals) |
| Treatment Strategy | - Randomization at end of run-in period<br><br>- Control arm: Lift policies immediately (0 days; cycle phase 3)<br>- Treatment arm: Maintain policies for full 90 days (cycle phase 2)<br><br>- Required adherence to assigned strategy for entire exposure period | - Identified states entering trials when transitioning from "high" to "low or declining" pandemic severity using NYTCDR<br>- Measured actual days of policy maintenance during 90-day exposure window or until "high" pandemic severity returns<br>- Used continuous PolicyDays variable (0-90) to emulate the binary comparison |
| Intervention Assignment | Random assignment to maintain policies for 0 days versus 90 days upon transitioning to "low/declining" severity periods | Confounding control using:<br>- State-level sociodemographic characteristics (% bachelors+; % homeownership)<br>- Time-varying confounding (days from pandemic start i.e. March 13, 2020)<br>- Inverse probability weighting for censoring |
| Follow-up | 150-day total observation window:<br>- 30-day run-in period<br>- 90-day intervention period<br>- 30-day follow-up period | Same; with censoring if:<br>- Return to "high" pandemic severity during exposure window<br>- Policy reinstatement during exposure window (States can re-enter trials at future time points) |
| Outcomes | State-level psychological distress prevalence change from run-in to follow-up periods (i.e. Distress_RunIn - Distress_FollowUp) | Change in psychological distress prevalence from 30-day run-in period to 30-day follow-up period<br>Measured using BRFSS as weighted proportion of positive mental health screens (>14 days criterion) |
| Causal Contrast | Intention-to-treat effect comparing depression prevalence change between 0-day and 90-day extended policy groups | Average treatment of 0 days versus 90 days of extended policy |
| Statistical Analysis | Two-sample comparison of depression prevalence change between randomized groups | Parametric G-computation with inverse probability weighting for censoring |
